# Pathogenic *SPTBN1* variants cause a novel autosomal dominant neurodevelopmental syndrome

**DOI:** 10.1101/2020.08.31.20184481

**Authors:** Margot A. Cousin, Keith A. Breau, Blake A. Creighton, Rebecca C. Spillmann, Erin Torti, Sruthi Dontu, Swarnendu Tripathi, Deepa Ajit, Kathryn M. Harper, Michael C. Stankewich, Richard E. Person, Yue Si, Elizabeth A. Normand, Amy Blevins, Alison S. May, Louise Bier, Vimla Aggarwal, Grazia M. S. Mancini, Marjon A. van Slegtenhorst, Kirsten Cremer, Jessica Becker, Hartmut Engels, Stefan Aretz, Jennifer J. MacKenzie, Eva Brilstra, Koen L. I. van Gassen, Richard H. van Jaarsveld, Renske Oegema, Gretchen M. Parsons, Paul Mark, Ingo Helbig, Sarah E. McKeown, Robert Stratton, Benjamin Cogne, Bertrand Isidor, Pilar Cacheiro, Damian Smedley, Helen V. Firth, Tatjana Bierhals, Katja Kloth, Deike Weiss, Cecilia Fairley, Joseph T. Shieh, Amy Kritzer, Parul Jayakar, Evangeline Kurtz-Nelson, Raphael Bernier, Tianyun Wang, Evan E. Eichler, Ingrid M.B.H. van de Laar, Allyn McConkie-Rosell, Marie McDonald, Jennifer Kemppainen, Brendan C. Lanpher, Laura E. Schultz-Rogers, Lauren B. Gunderson, Pavel N. Pichurin, Grace Yoon, Michael Zech, Robert Jech, Juliane Winkelmann, Undiagnosed Diseases Network, Genomics England Research Consortium, Michael T. Zimmermann, Brenda Temple, Sheryl S. Moy, Eric W. Klee, Queenie K.-G. Tan, Damaris N. Lorenzo

## Abstract

*SPTBN1* encodes βII-spectrin, the ubiquitously expressed member of the β-spectrin family that forms micrometer-scale networks associated with plasma membranes. βII-spectrin is abundantly expressed in the brain, where it is essential for neuronal development and connectivity. Mice deficient in neuronal βII-spectrin expression have defects in cortical organization, global developmental delay, dysmorphisms, and behavioral deficiencies of corresponding severity. These phenotypes, while less severe, are observed in haploinsufficient animals, suggesting that individuals carrying heterozygous variants in this gene may also present with measurable compromise of neural development and function. Here we report the identification of heterozygous *SPTBN1* variants in 29 individuals who present with global developmental, language and motor delays, mild to severe intellectual disability, autistic features, seizures, behavioral and movement abnormalities, hypotonia, and variable dysmorphic facial features. We show that these *SPTBN1* variants lead to loss-of-function, gain-of-function, and dominant negative effects that affect protein stability, disrupt binding to key protein partners, and disturb cytoskeleton organization and dynamics. Our studies define the genetic basis of this new neurodevelopmental syndrome, expand the set of spectrinopathies affecting the brain and neural development, and underscore the critical role of βII-spectrin in the central nervous system.

## Introduction

Spectrins are ubiquitously expressed, elongated polypeptides that bind membrane lipids and ankyrins to line the plasma membrane^1,2^. The spectrin meshwork is formed by heterodimeric units of α- and β-spectrin assembled side-to-side in antiparallel fashion, which then form head-to-head tetramers that crosslink F-actin to form spectrin-actin arrays^1,2^. Mammalian neurons express the most diverse repertoire of spectrins (αII- and βI-V spectrins) of any cell type^3^. Together with ankyrins, spectrins self-assemble with both remarkable long-range regularity and micro- and nanoscale specificity to precisely position and stabilize cell adhesion molecules, membrane transporters, ion channels, and other cytoskeletal proteins^3^. Some spectrins also enable intracellular organelle transport^3^. Unsurprisingly, deficits in spectrins underlie several neurodevelopmental and neurodegenerative disorders^4-6^. For example, inherited autosomal dominant variants in Plll-spectrin (encoded by *SPTBN2)* cause late onset spinocerebellar ataxia type 5 (SCA5)^5^, while pathogenic *de novo* variants have been associated with early childhood ataxia, intellectual disability (ID), and developmental delay (DD)^7-12^. Similarly, autosomal recessive *SPTBN2* variants^13-15^ are associated with childhood ataxia, which may also present with ID and DD^13^, collectively referred to as autosomal recessive spinocerebellar ataxia 14 (SCAR14). *De novo* pathogenic variants in *SPTAN1*, which encodes αII-spectrin, cause West syndrome, an early-infantile epileptic encephalopathy (EIEE) characterized by frequent severe seizures and persistent abnormality of cortical function^5^, and other childhood onset epileptic syndromes^16-20^. Some patients co-present with spastic quadriplegia, DD, and various brain defects^5^. In addition, dominantly inherited *SPTAN1* nonsense variants were recently linked to juvenile onset hereditary motor neuropathy^21^. Biallelic alterations in βIV-spectrin (encoded by *SPTBN4)* result in congenital hypotonia, neuropathy, and deafness, with and without ID^6,22,23^.

Neuronal βII-spectrin, encoded by *SPTBN1*, is the most abundant β-spectrin in the brain and forms tetramers with αII-spectrin, which intercalate F-actin rings to build a sub-membranous periodic skeleton (MPS)^24^. A cytosolic pool of βII-spectrin promotes bidirectional axonal organelle transport^25,26^. We previously reported that mice lacking βII-spectrin in all neural progenitors *(Sptbn1^flox/flox^*;Nestin-Cre; referred to as βIISp-KO) show early postnatal lethality, reduced long-range cortical and cerebellar connectivity, spontaneous seizures, and motor deficits^26^. However, the impact of human genetic variation in *SPTBN1* on βII-spectrin function and its association with disease has not been studied. Here we describe a cohort of 29 individuals carrying rare, mostly *de novo* variants in *SPTBN1* affected by a novel autosomal dominant neurologic syndrome presenting with global developmental, language and motor delays, mild to severe ID, autistic features, seizures, behavioral abnormalities, hypotonia, and variable dysmorphisms. This suggests conserved roles for βII-spectrin in neuronal development and function. The most damaging variants clustered within the actin-binding calponin homology domain (CH) and led to aberrant neuronal morphology, decreased neurite outgrowth, and deficient axonal organelle transport in primary neurons. Consistent with these deficiencies, our biochemistry, microscopy, and molecular modeling studies indicate that *SPTBN1* variants lead to loss-of-function (LOF), gain-of-function (GOF), and dominant negative effects that affect protein stability, disrupt binding to key protein partners, and affect cytoskeleton organization and dynamics. Consequently, histology and behavioral studies in brain βII-spectrin-deficient mice showed neuron-autonomous brain connectivity defects and recapitulated developmental and behavioral phenotypes observed in patients with *SPTBN1* variants. Collectively, our data strongly support pathogenic mechanisms of *SPTBN1* variants as the genetic cause of a novel neurodevelopmental syndrome and underscores the multifaceted role of βII-spectrin in the nervous system.

## Results

### Patients with *SPTBN1* variants present with a novel neurodevelopmental syndrome

A cohort of 29 individuals from 28 families (one pair of siblings) who carry heterozygous variants in *SPTBN1* was identified through whole genome (WGS) or exome (WES) sequencing. These probands presented with neurodevelopmental delay and variable neurologic, behavioral, and dysmorphic features (Fig. 1, Table 1, Supplementary Note). Twenty-four of the 29 affected individuals carry *de novo* variants, while the remaining have unknown inheritance due to lack of parental samples for testing (Supplementary Table 1). In proband P17 the *SPTBN1* variant inheritance was unknown but sequencing revealed mosaicism at 13.3% of reads suggesting the variant occurred *de novo*. Twenty-eight unique variants are described (P10 has two *de novo* variants in cis) of which 22 are missense, three are nonsense, and three are canonical splice-site variants, with two predicted by SpliceAI^27^ to lead to in-frame deletions and one predicted to result in a frameshift that introduces a premature stop codon (Fig. 1a, Supplementary Table 1). Approximately half of the variants cluster in the CH domain, predominantly in the second CH domain (CH2), with the rest distributed in various spectrin repeats (SR) (Fig. 1a).

**Fig. 1:**
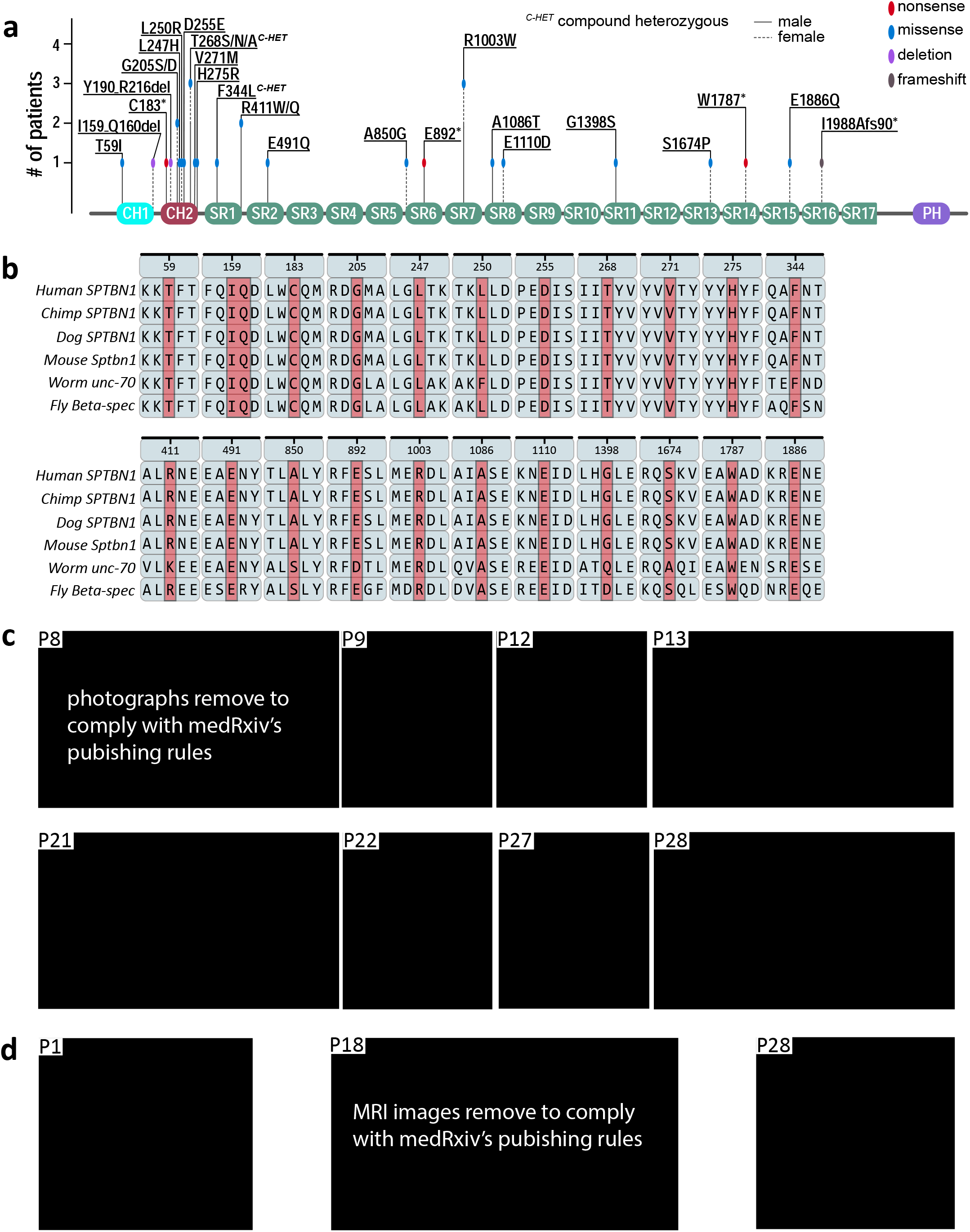
*SPTBN1* variants found in individuals with neurodevelopmental disorders. **a**, Schematic representation of functional domains of βII-spectrin. CH1=calponin homology domain 1 (teal), CH2=calponin homology domain 2 (red), SR=spectrin repeat (green), and PH=pleckstrin homology domain (purple). The locations of *SPTBN1* variants are indicated. **b**, Alignment of protein sequences for βII-spectrin and orthologues show that missense variants identified in the patients in this study are located at highly conserved residues across species from humans to *Drosophila*. The position of *SPTBN1* variants analyzed in the sequenced of human βII-spectrin is shown for reference. **c**, Photos of individuals with *SPTBN1* variants. Ages at the time of photograph are: P8: 7y8m, P9:16, P12: 11y, P13: 6y, P21 left: unknown, Right: 11y, P22: 15y, P27: 16y11m, P28: 3y11m. **d**, Examples of brain MRI findings: diffuse cerebral parenchymal volume loss (L>R) and asymmetric appearance of hippocampi (P1, acquired at <1y), white matter disease in the supratentorial and infratentorial regions (P18, acquired at 7y), thinning of the posterior body of the corpus callosum without significant volume loss (P28, acquired at 10m).

**Table 1.**
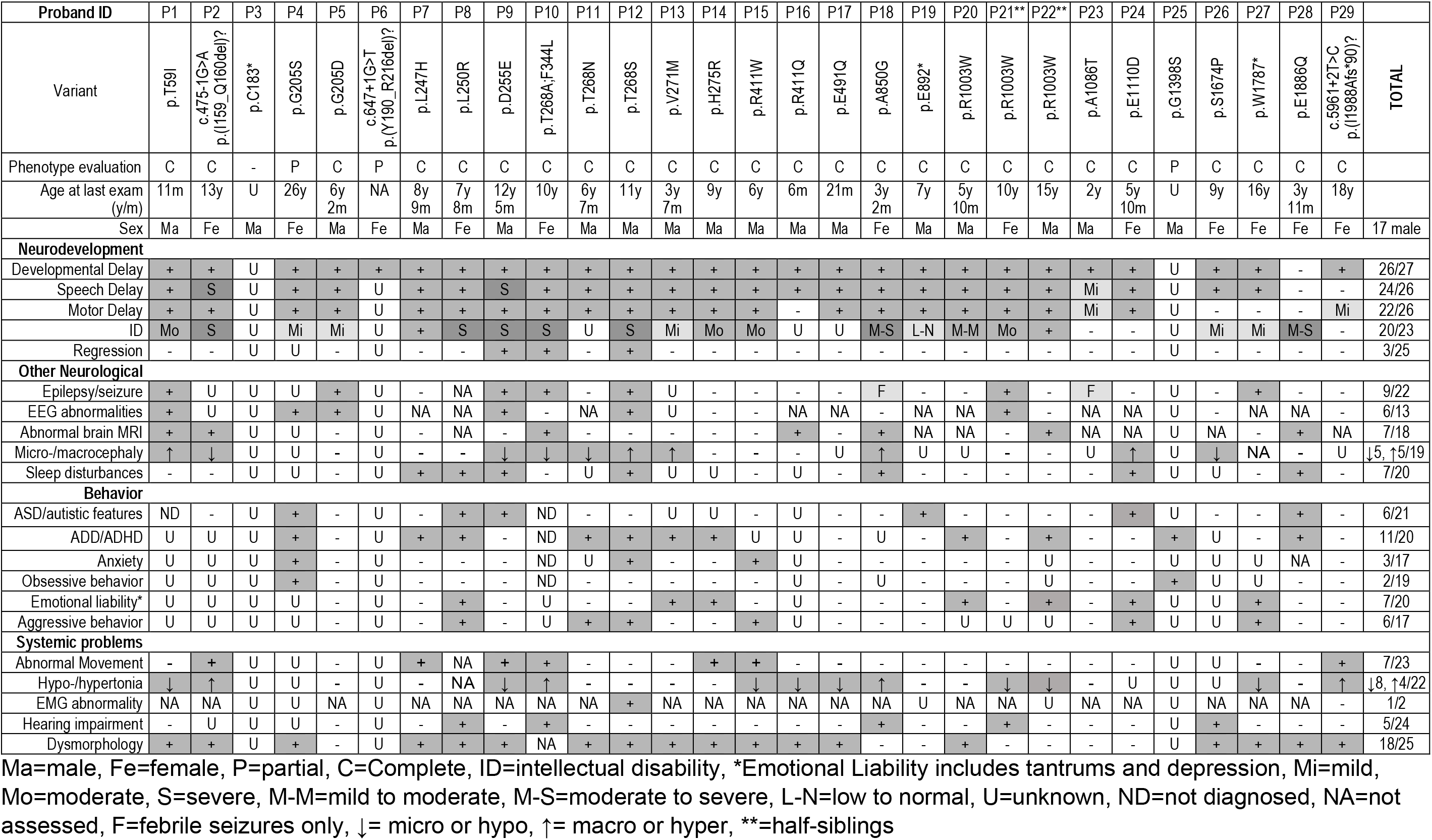
Summary of clinical features observed among *SPTBN1* mutations carriers. Abbreviations are as follows: Ma=male, Female=F, P=partial, C=complete, ID=intellectual disability, *Emotional Liability includes tantrums and depression, Mi=mild, Mo=moderate, S=severe, M-M=mild to moderate, M-S=moderate to severe, L-N=low to normal, U=unknown, ND=not diagnosed, NA=not assessed, F=febrile seizures only, +=presence, -=absence, ↓= micro or hypo, ↑= macro or hyper, **=half-siblings.

The phenotypic findings are summarized in Table 1 and detailed clinical and family histories are included in the Supplementary Note. The cohort included 17 male and 12 female probands with the age at last evaluation spanning from 6 months to 26 years of age. All had early onset of symptoms with primarily developmental delay (DD) presentations with the exception of two individuals. For 28 individuals with phenotypic data, 26 reported some level of delays. Of those with DD, 21 reported both speech (SD) and motor (MD) delays, three reported only SD (P16, P26 and P27), and one reported only MD (P29). Twenty individuals reported ID, with seven of those being severe or moderate to severe, and six being mild or low to normal. Developmental regression was noted in three individuals. Although mild delays were noted in the clinical evaluation of proband P29 (c.5961+2T>C, p.(I1988Afs*90)), the primary symptom, dystonia, was observed at age 13. Similarly, while delay in speaking was noted for proband P23 (p.A1086T), the primary phenotype in this individual was liver-related. Only partial phenotype information was obtained for previously reported probands P4 and P25^28^, and no specific phenotypic information is available for P3 from the Deciphering Developmental Disorders (DECIPHER) database^29^, although this individual was presumed to have some form of DD (Supplementary Note).

Nine individuals have a history of seizures, with two being febrile only, and six with electroencephalogram (EEG) correlates. Seizure episodes in four of the cases were resolved later in childhood or were successfully managed through medication. Four of these patients were diagnosed with frontal lobe or generalized epilepsy. Seven individuals had abnormal brain MRI findings, including three patients with thinning of the corpus callosum (CC), two with ventriculomegaly, two with delayed myelination, and two showing some volume loss (P1: diffuse cerebral parenchymal; P10: mild cerebellar and vermian). The remaining brain MRI findings were unique (Table 1, Supplementary Note). Behavioral concerns were common within the cohort. Six individuals displayed autistic features or had an autism spectrum disorder (ASD) diagnosis, including two (P19 and P24) previously reported as part of a WES study of a 2,500 ASD patient cohort^30^. One other proband (P25) was originally identified as part of a WES study of over 500 trios from Tourette syndrome cohorts^28^. Thirteen individuals presented with other behavioral concerns, including attention deficit and hyperactivity disorder (ADD/ADHD) (n=11), anxiety (n=3), emotional liability including tantrums and depression (n=7), and aggressive or self-injurious behaviors (n=6). Seven individuals experienced sleep disturbances, in some cases co-occurring with seizure episodes. Additional phenotypic findings include changes in muscle tone and movement abnormalities. Ten had hyper- (n=4) or hypotonia (n=8). Seven individuals reported movement abnormalities including dystonia (n=3), ataxic or unsteady gait (n=5), spasticity (n=1), and tremor (n=2). Less common features included hearing impairment, reported for five individuals, generally mild to moderate, with one individual suspected to have conductive hearing loss due to recurrent ear infections. Dysmorphic features were noted in 18 individuals with a subset of individuals shown in Fig. 1c, but no consistent findings were observed. Macrocephaly (n=5) and microcephaly (n=5) were both observed as well as other head shape anomalies (n=6).

*SPTBN1* is intolerant to both missense and loss-of-function variants (gnomAD v2.1.1)^31^, and protein sequence alignment of human βII-spectrin and its orthologues across several species shows a high degree of evolutionary conservation of the residues impacted by these putative pathogenic variants (Fig. 1b). Consistent with their implied functional relevance, the majority of the variants are predicted to be likely damaging to protein function by multiple prediction tools (PolyPhen-2, Mutation Taster, SIFT, PROVEAN, M-CAP, PredictSNP2, and CADD) (Supplementary Table 1). Additionally, all variants are absent or extremely rare in the population (gnomAD v2.1.1)^31^ (Supplementary Table 1). Missense variants in codons G205, T268, R411 and R1003 were identified in more than one individual (Fig. 1a). The p.R1003W variant was identified in two maternal half-siblings (P21 and P22) inherited from their unaffected mother, who was found to be mosaic for the variant at a low level (found in 1.8% of next generation sequencing reads). Unrelated individual P20 also carries the *de novo* p.R1003W variant, and has the common DD features, but also presented with some non-overlapping clinical features. Similarly, variants in unrelated duos P4 and P5, and P15 and P16 affect the same p.G205 and p.R411 residues, respectively, but result in different amino acid substitutions. All of these individuals have DD, both P4 and P5 had an abnormal EEG, and both P15 and P16 had hypotonia but each also has some distinct features consistent with the variability in the cohort. Likewise, unrelated patients P10, P11, and P12 carry different amino acid substitutions in residue p.T268, and present with overlapping phenotypes. Notably, P10 has two βII-spectrin variants *in cis* (p.F344L and p.T268A), which may contribute to the more severe phenotype observed. The partial clinical divergence within these patients likely stems from differences in sex, age, and genomic background, which in turn may determine their corresponding penetrance and physiological consequences.

In sum, the above clinical presentations suggest that *SPTBN1* variants converge to impair cellular and physiological mechanisms that lead to delays in motor and language development and cognitive skills. Additionally, the results of these evaluations suggest that several of these variants also result in additional neurological and behavioral phenotypes. These observations are consistent with pleiotropic functions of βII-spectrin including its diverse and critical roles in brain development and function^26^.

### Human βII-spectrin mutations affect protein cellular distribution and alter cell morphology

To begin to assess the pathogenic mechanisms of *SPTBN1* variants, we introduced a subset of the mutations in the coding sequence of human βII-spectrin cloned into the peGFP-C3 plasmid, transfected the constructs into HEK293 cells, either alone or together with pmCherry-C1, and monitored their effects on GFP-βII-spectrin (GFP-βIISp) levels, localization, size, and stability by confocal microscopy and western blot. Of the 22 mutations tested, protein levels, of 10 were unchanged relative to control (Fig. 2a, Extended Data Fig. 1a). Variants p.H275R and p.A850G resulted in 25% and 50% βII-spectrin overexpression, respectively. Expression of nonsense p.C183*, p.E892* and p.W1787* variants yielded truncated proteins of the expected size. However, while p.W1787* was expressed normally, the levels of the p.C183* and p.E892* mutants were significantly reduced. While neither of the two CH domain in-frame deletions (I159_Q160del and Y190_R216del) affected protein levels, several mutations in this region showed lower expression. Both p.G205D and p.G205S mutations reduced protein levels, with p.G205D also impacting βII-spectrin solubility, which largely precipitated into the Triton-X100 insoluble fraction (Fig. 2b, Extended Data Fig. 1b). This indicates that these amino acid substitutions at p.G205 likely affect the structural conformation of βII-spectrin, and result in unfolded and unstable polypeptides. Notably, wildtype (WT) GFP-βIISp localized through the cytosol and at the cell membrane whereas GFP-βIISp bearing p.l159_Q160del at the end of CH1, or the p.C183*, p.Y190_R216del, p.G205D, and p.G205S mutations in the proximal CH2 domain accumulated in large cytosolic aggregates (white arrowheads Fig. 2c and Extended Data Fig. 1c). Expression of the CH2 mutations p.T268A, T268N, T268S, V271M, and H275R resulted in normal GFP-βIISp distribution, but induced noticeable changes in cell morphology. Overall, cells were enlarged and had increased membrane protrusions (Fig. 2c and Extended Data Fig. 1c, asterisks). These changes are indicative of modified cytoskeleton arrangements and dynamics, and likely reflect altered F-actin binding.

**Fig. 2:**
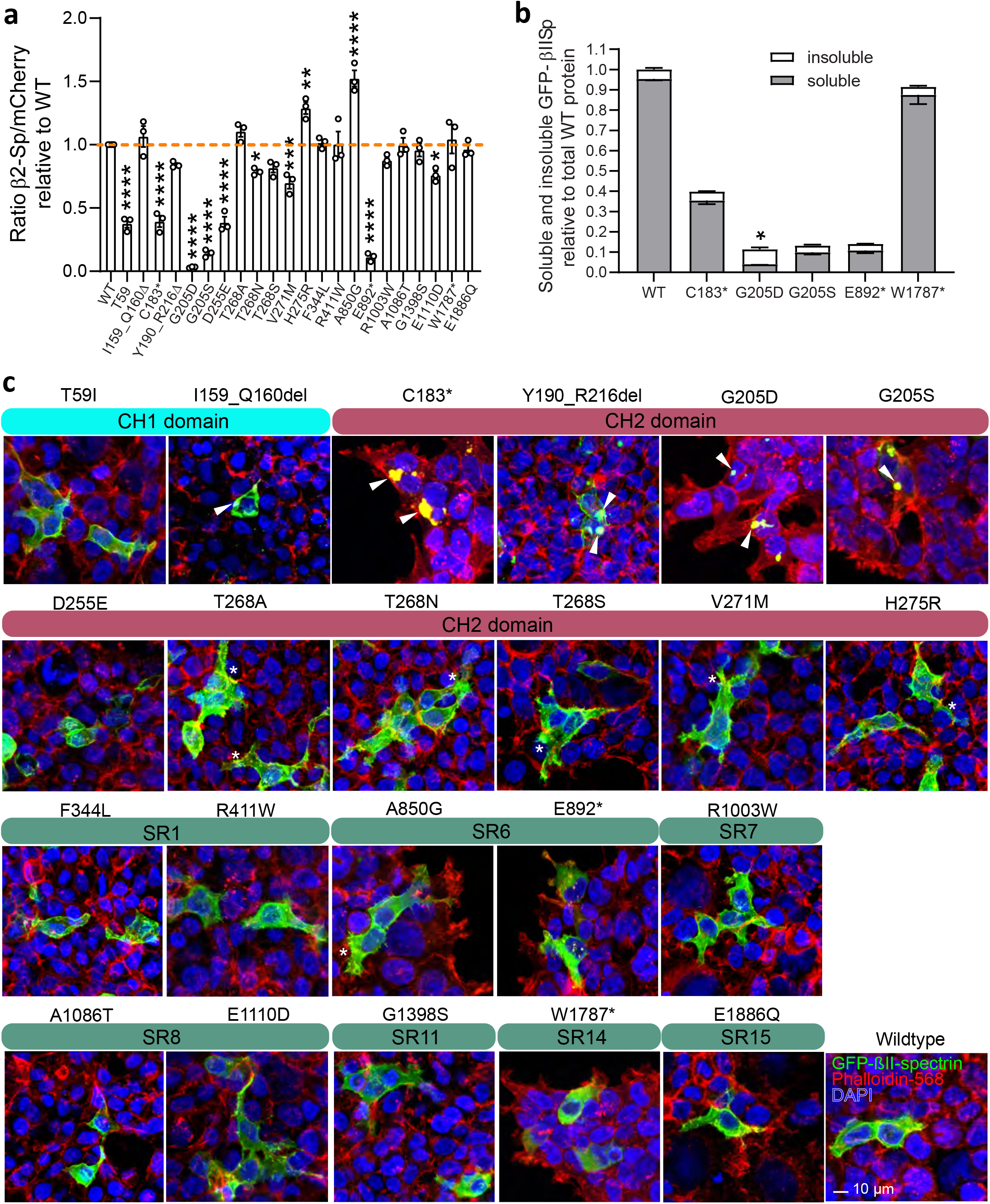
*SPTBN1* variants alter protein expression and subcellular distribution. **a**, Levels of GFP-βIISp mutant proteins in HEK293T cells co-transfected with GFP-βIISp and mCherry plasmids relative to expression of WT GFP-βIISp. **b**, Partition of indicated GFP-βIISp proteins expressed in HEK293T cells between Triton-X100 soluble and insoluble fractions relative to total WT GFP-βIISp levels. Data in b and c is representative of three independent experiments. All data represent mean ± SEM. One-way ANOVA with Dunnett’s post hoc analysis test for multiple comparisons, *p < 0.05, ** p< 0.01, ***p < 0.001, ****p < 0.0001. **c**, Immunofluorescence images of HEK293T cells transfected with indicated GFP-βIISp plasmids and stained for actin (phalloidin) and DAPI. Scale bar, 10 μm. Expression of variants within the distal end of CH1 and the proximal portion of the CH2 domains result in cytosolic GFP-positive aggregates (white arrowheads). Expression of variants within the C-terminal portion of the CH2 domain and a subset of variants in SRs increase the number of membrane protrusions (white asterisks). Data is representative of at least six independent experiments.

Cell morphology phenotypes induced by SR mutations varied. For example, cells expressing the p.R411W mutation, located in SR1, which is required for dimerization with αII-spectrin^32^ and thought to contribute to actin binding^33^, were enlarged and with increased membrane protrusions. Among the five mutations clustered within SR6-8, p.A850G and p.R1003W induced the most noticeable morphological changes (Fig. 2c, Extended Data Fig. 1c). Interestingly, p.A850G, which results in βII-spectrin overexpression, also causes a striking increase in membrane protrusions, which suggests a GOF effect (Fig. 2c and Extended Data Fig. 1c). Since SR3-S14 have no assigned functional specificity nor contain known binding sites for partners, a molecular rationale for how mutations in this region affect βII-spectrin function is lacking. SR15 binds ankyrins, with amino acid p.Y1874 known to be critical for this interaction^34^. However, p.E1886Q GFP-βIISp does not show apparent cellular changes, likely because it does not disrupt binding to ankyrins^34^, or this complex is not required for cytoskeleton organization or dynamics in HEK293 cells. Expression of E892* and W1787* GFP-βIISp, lacking the polypeptide portions from SR6 to C-terminus and SR14 to C-terminus respectively, did not cause apparent cellular phenotypes. This is surprising because in addition to loss of ankyrin binding, these truncated proteins also lack the tetramerization^35^ and pleckstrin homology (PH) domains^36^, the latter being important for binding lipids in the cell and organelle membranes. Together, these data indicate that human βII-spectrin mutations can lead to cellular phenotypes through LOF and GOF mechanisms that likely involve changes in cytoskeleton architecture and dynamics.

### Human βII-spectrin mutations affect its interaction with submembrane cytoskeleton partners

A subcortical network of F-actin- and ankyrin-bound βII-/αII-spectrin tetramers promotes membrane stability and helps organizing membrane proteins within specialized microdomains^1-3^. Thus, pathogenic *SPTBN1* variants could impair neuronal development and/or function by altering βII-spectrin interaction with F-actin and other cytoskeletal partners or their submembrane availability. Consistent with the latter prediction, we found that both actin and mCherry-αIISp were sequestered in GFP-βIISp aggregates caused by expression of various CH domain variants in HEK293 cells (arrowheads, Fig. 3a, Extended Data Fig. 2a). These mutations in the CH domain (F-actin binding region) also resulted in GFP-βIISp aggregation and in clustering of endogenous actin and αII-spectrin within GFP aggregates when expressed in cortical neurons from βII-SpKO mice (arrowheads, Fig. 3b). To further evaluate the ability of mutant βII-spectrin to associate with molecular partners, we conducted binding assays and co-immunoprecipitation (co-IP) experiments. We prioritized the evaluation of a subset of variants based on the likelihood that they would have an effect on the interaction tested, given their position on the specific domains known to be critical for binding to that specific partner. We first assessed the effect of the mutations on the formation of βII-spectrin/αII-spectrin complexes by incubating GFP beads coupled to WT or mutant GFP-βIISp with cell lysates expressing mCherry-αIISp and measuring the amount of mCherry-αIISp in eluates from GFP pulldowns by western blot. As expected, mutant C183* GFP-βIISp neither associated with mCherry-αIISp in pulldown assays nor sequestered mCherry-αIISp or endogenous αII-spectrin into GFP-βIISp aggregates in HEK293 or neurons because it lacks the heterodimer nucleation SR1-SR2 region^30^ (Fig. 3a-c and Extended Data Fig. 2b). Similarly, the pulldown of αII-spectrin with G205D and G205S GFP-βIISp baits yielded less αII-/βII-spectrin complexes, partly due to the lower expression of these mutant polypeptides, but it also indicates a lower affinity for αII-spectrin (Fig. 3a-c and Extended Data Fig. 2b). That a single substitution in CH2 reduces αII-spectrin affinity is surprising because this domain has not been linked to αII-spectrin binding. With the exception of the p.R1003W mutant located in SR7, which reduces association with αII-spectrin by 40%, none of the other variants tested affects αII-spectrin binding (Fig. 3c and Extended Data Fig. 2a, b). The lower αII-spectrin binding to R1003W GFP-βIISp might result from local or long-range conformational changes that weakens interactions along the dimer.

**Fig. 3:**
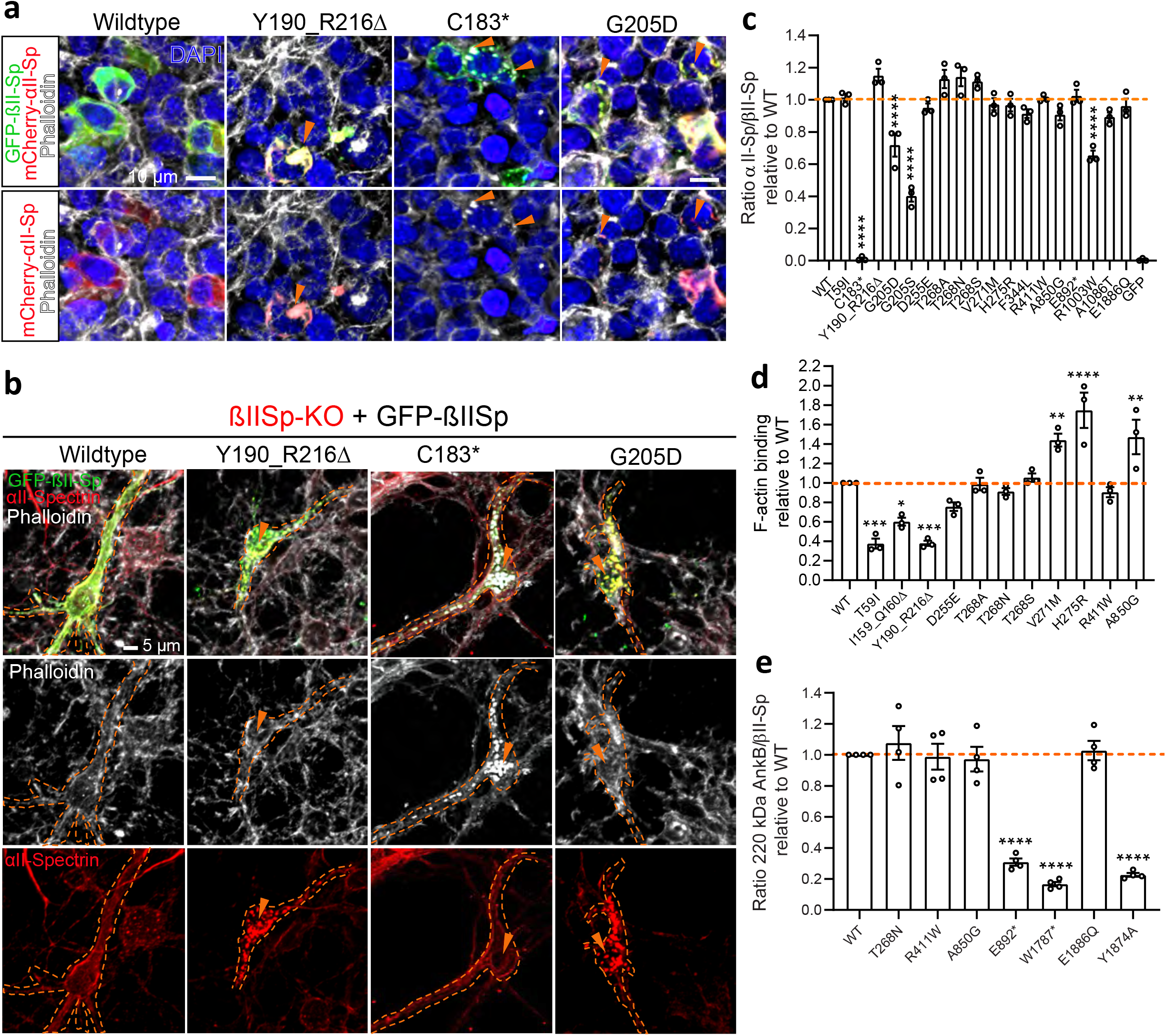
*SPTBN1* variants alter interaction with critical cytoskeleton partners. **a**, Immunofluorescence images of HEK293T cells transfected with mCherry-αIISp and with either WT or mutant GFP-βIISp plasmids. Cells were stained for actin (phalloidin) and DAPI. Scale bar, 10 μm. **b**, Immunofluorescence images of DIV8 mouse βIISp-KO cortical neurons transfected with indicated GFP-βIISp plasmids and stained for actin (phalloidin) and endogenous αII-spectrin. Scale bar, 5 μm. in a and b GFP-positive cytoplasmic aggregates (orange arrowheads) also contain either actin or αII-spectrin proteins, or both. **c**, Quantification of binding of mCherry-αIISp to GFP-βIISp proteins relative to the abundance of mCherry-αIISp/WT GFP-βIISp complexes. **d**, Binding of purified βII-spectrin proteins to purified F-actin assessed through an actin co-sedimentation assay. **e**, Binding of GFP-βIISp proteins to 220-kDa AnkB-2HA assessed via co-IP from HEK293T cells. The Y1874A βII-spectrin mutation known to disrupt the formation of AnkB/ βIISp complexes was used as control. Graphs in c and d summarize results from three independent experiments. Data in e summarizes four independent experiments. All data represent mean ± SEM. One-way ANOVA with Dunnett’s post hoc analysis test for multiple comparisons, *p < 0.05, ** p< 0.01, ***p <0.001, ****p < 0.0001.

Next, we evaluated whether mutations in the CH domains, the known actin-binding domain in βII-spectrin, affect its binding to F-actin using a co-sedimentation assay. βII-spectrin proteins containing a PreScission protease (PP) recognition site between GFP and the initiation codon of βII-spectrin (GFP-PP-βIISp) were produced in HEK293 cells and captured on GFP beads. Purified WT and mutant βII-spectrin were recovered from beads upon PP cleavage and mixed with purified F-actin. The partition of βII-spectrin between the soluble (S) and actin-containing pellet (P) fractions was used to estimate the relative binding proclivity between both proteins. We found that CH1 mutation p.T59I and deletion p.I159_Q160del in the CH1-CH2 linker led to approximately 40% and 70% less βII-spectrin association with F-actin, respectively (Fig. 3d and Extended Data Fig. 2c). Similarly, p.Y190_R216del and p.D255E CH2 variants also resulted in reduced F-actin binding. In contrast, the p.V271M and p.H275R mutations increased F-actin binding to βII-spectrin by 60-90%, while all three p.T268S/N/A mutants bound F-actin at similar levels as the WT protein (Fig. 3d and Extended Data Fig. 2c). This range in binding affinity is likely caused by the balance of individual or combined effects of both local and CH domain-wide conformational changes caused by modified intramolecular interactions, which in turn results in modified intermolecular contacts at the βII-spectrin/F-actin interface. Given that the p.A850G mutant causes a cell morphology phenotype similar to the ones induced by some of the CH domain mutants (Fig. 2c and Extended Data Fig. 1c), we also tested its binding to F-actin. Surprisingly, this mutant resulted in approximately 50% higher F-actin binding (Fig. 3d and Extended Data Fig. 2c). While this higher propensity for actin binding is likely to underlie the abnormal cell morphology, it is not clear how this substitution, several SR away from the CH domain, can modify this interaction.

Finally, we evaluated the impact of βII-spectrin mutations on its interaction with ankyrins. In this experiment, HA-tagged 220kDa ankyrin-B was expressed in HEK293 cells together with WT or mutant GFP-βIISp proteins. The presence of HA signal in eluates from GFP-βIISp complexes was detected by western blot. Consistent with previous reports^34^, expression of the Y1874 mutation in SR15 (the known ankyrin-binding domain) almost entirely abrogated binding to ankyrin-B (Fig. 3e and Extended Data Fig. 2d). As expected, truncated βII-spectrin polypeptides that lack SR15 caused by p.E892* and p.W1787* variants also disrupted binding between these partners (Fig. 3e and Extended Data Fig. 2d). Interestingly, the SR15 p.E1886Q variant did not affect binding to ankyrin-B, despite its spatial proximity to the Y1874 binding site^34^.

### Molecular modeling predicts effects of βII-spectrin variants on protein stability and F-actin binding

We further assessed the impact of *SPTNB1* variants through molecular modelling. We first modeled the 10 missense variants involving seven residues in the βII-spectrin CH1-CH2 domain. The CH domain is a protein module of around 100 residues composed of four alpha helices^37^ found in cytoskeletal and signal transduction actin-binding proteins (ABP)^38^. Multiple biochemical studies using ABP containing CH1-CH2 domains, such as spectrin superfamily members α-actinin-4 *(ACTN4)* and utrophin *(UTRN)*, suggest dynamic transitions between “closed” and “open” configurations of the tandem domains, whereas the open state is thought to expose CH1 residues to enable its predominant role of binding actin, with CH2 regulating the conformational state through autoinhibition^38^. The electrostatic surface profile of βII-spectrin CH1 and CH2 domains modeled using an available crystal structure of utrophin^39^ indicates that they each have one electrically active side complementary to each other and one neutral side, consistent with an energetically balanced closed conformation (Fig. 4a-c). This model also indicates that six of the eight mutated CH domain residues reside at the CH1-CH2 dimer interface, potentially impacting interdomain helix-helix interactions, thereby dysregulating the natural autoinhibition (Fig. 4b,c).

**Fig. 4:**
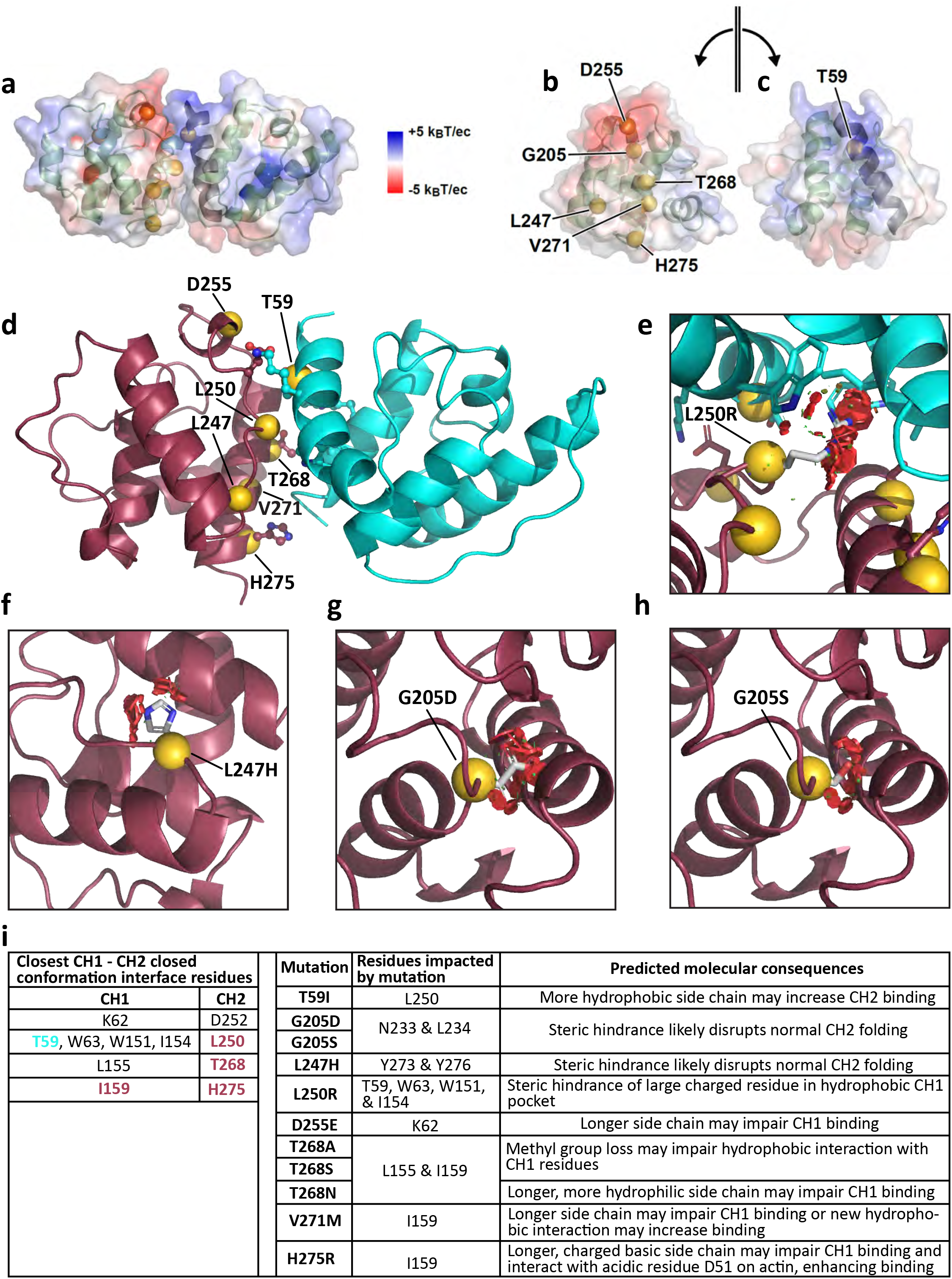
βII-spectrin CH domain variants likely alter CH1-CH2 dimer stability. **a**, Closed conformation of the βII-spectrin CH1-CH2 dimer modeled after utrophin^37^ showing the sites of βII-spectrin variants at the interface and the electrostatic surface of each domain calculated independently. **b, c**, Electrostatic complementarity shows that both CH domains have a polar side, where CH2 is negatively charged (b) and CH1 is positively charged(c), and both have a neutral side. **d**, Closed conformation of the βII-spectrin CH1-CH2 dimer modeled by docking the CH2 domain of βII-spectrin^38^ onto the CH1 domain modeled after PIII-spectrin^39^. **e**, The L250R variant introduces a large, positively charged residue that clashes with a hydrophobic CH1 pocket through steric hindrance and electric instability. **f**, L247H introduces a large aromatic amino acid and likely disrupts normal CH2 folding. **g, h** Steric hindrance and negative charge introduced by (g) G205D and (h) G205S in the interior of CH2 likely disrupts normal CH2 folding. **i**, Key interactions at the CH1-CH2 interface (site of mutations in CH1 (teal) and CH2 (red)) and likely molecular perturbations caused by *STPBN1* variants.

To refine our prediction of the closed conformation of the βII-spectrin CH1-CH2 domain and to identify interactions at the interface, we docked the CH2 domain (residues 173-278) of βII-spectrin^40^ onto the available modeled structure of the CH1 domain (residues 55-158) of pill-spectrin (95% homologous with βII-spectrin)^41^ and chose the top docking pose (Fig. 4d). This pose was also the same pose compared to the actinin-4 (PDB ID 6oa6) (unpublished) and utrophin (PDB ID 1qag)^39^ closed conformations (Extended Data Fig. 3a-c). Figure 4i summarizes the key predicted interacting residues at the CH1-CH2 interface and the structural consequences of the variants in those domains. Four of the eight residues affected by *SPTBN1* missense variants (T59 in CH1, and L250, T268, and H275 in CH2) are predicted to be involved in interdomain interactions (Fig. 4d,i). The *SPTBN1* missense variants in these and the two other interface residues (D255 and V271) are predicted to introduce destabilizing effects (Fig. 4i). For example, substitutions of T268 by Ala (smaller and more hydrophobic), Ser (loss of methyl group), and Asn (larger and more hydrophilic) likely have different degrees of alteration of the original T268 hydrophobic interaction with L155 in CH1 and I159 in the CH1-CH2 linker. However, if these amino acid changes in T268 affect the CH1-CH2 domain conformation, they do not result in appreciable changes in F-actin binding (Fig. 3d and Extended Data Fig. 2c), despite their marked cellular (Fig. 2c, Extended Data Fig. 1c) and disease-linked effects. Similarly, substitution of glutamic for aspartic acid in D255E is a relatively small change that does not result in changes in F-actin affinity. On the other hand, both the V271M (larger and hydrophobic) and the H275R (longer and significantly more hydrophilic) substitutions may impair CH1 binding to cause a shift toward the open CH1-CH2 conformation and higher F-actin affinity. This is also likely the case for the L250R mutation, which is expected to cause significant steric hindrance by the clashing of the large, charged residue with a hydrophobic CH1 pocket (Fig. 4e). The molecular and cellular consequences of this variant remain to be assessed. Conversely, the T59I mutation introduces a slightly longer, but more hydrophobic group that might promote a stronger interaction with L250 in CH2, potentially shifting the equilibrium to a CH1-CH2 closed configuration consistent with less F-actin binding (Fig. 3d and Extended Data Fig. 2c).

The amino acid substitutions in the two CH2 sites in the interior of the domain (G205S/D and L247H) are predicted to cause significant steric hindrance, which likely results in CH2 domain instability (Fig. 4f-h). G205S/D introduces destabilization by positioning a negative charge on the interior and steric hindrance against the neighbor N233 and L234 sidechains (Fig. 4g,h), which likely underlies the aggregation of the mutant protein in cells (Figs. 2c and 3b). The Y190_R216del mutation, which eliminates 27 amino acids in the CH2 domain, also results in βII-spectrin aggregation and diminished F-actin binding (Figs. 2c and 3b,d and Extended Data Fig. 2c). In these cases, the autoinhibitory interactions will also be lost if the structure of the CH2 domain is compromised, but through a different mechanism than the genetic variants that alter the CH dimer interface. Besides the open/closed CH1-CH2 domain conformational shifts, some of these mutants might be directly involved in binding F-actin. To explore this possibility, we independently docked the CH1 and CH2 domains onto an F-actin model built from chains A-F of 6anu^40^ using ClusPro^42,43^ (Extended Data Fig. 3d-f). The top eight CH1 docking poses predicted by the balanced and electrostatic scoring algorithms almost all correspond to the location and orientation of CH1 molecules on F-actin as defined by the cryo-EM structure 6anu (Extended Data Fig. 3d, dark blue). For CH2 docking onto F-actin, the top eight docking poses predicted by the balanced and electrostatic scoring algorithms almost all correspond to symmetry-related locations and poses on F-actin (Extended Data Fig. 3e). In addition, the predicted orientation of CH2 molecules on F-actin is consistent with the known binding site of the CH1 domain, as judged by the length of the linker that would be required to join the C-terminus of the docked CH1 domain to the N-terminus of the docked CH2 domain (Extended Data Fig. 3f). Our model predicts that neither the T59 residue nor its mutated version are directly involved in F-actin binding (Extended Data Fig. 3d). On the CH2 domain, the H275R mutant may result in a stronger interaction with negatively charged D51 in F-actin (Extended Data Fig. 3e), which may further contribute to its higher actin binding propensity (Fig. 3d and Extended Data Fig. 2c). We also modeled the missense mutations in the SR (Extended Data Fig. 3g,h). Except for F344L, all SR variants face outwards, to the solvent, indicating that they could be involved in protein binding at the interface. Interestingly, all mutations within the second and third helices of the spectrin fold are neutral or more hydrophobic for the variants, and those in the first helix of the SR are more hydrophilic. Given the consistency of this trend, we suspect that it may underlie a conserved functional role important for heterodimerization and larger order assemblies.

In sum, our modeling results provide a strong molecular rationale for the biochemical and cellular observations described above, which implicate protein stability, abnormal assembly and dynamics of the βII-spectrin-F-actin skeleton, and potential disruptions of βII-spectrin binding to other molecular partners, consistent with similar LOF and GOF changes observed in other members of the spectrin superfamily^37,40,43^.

### βII-spectrin mutations disrupt neuron architecture and function

Individual with *SPTBN1* variants display developmental deficits and a wide range of neurological phenotypes, which implicate βII-spectrin in neuronal development and cerebral cortex function. These clinical presentations are consistent with phenotypes of neural progenitor-specific βII-spectrin null mice that lack βII-spectrin throughout brain development^26^. In addition, cortical and hippocampal neurons from these mice show disruption of the spectrin-actin membrane periodic structure^24^, impaired axonal formation and growth, and reduced axonal organelle transport, all deficits that can be rescued by expression of WT βII-spectrin^26,44^. These reports, together with our initial cellular and molecular observations shown above, suggest that mutant βII-spectrin may result in defects in the organization and the dynamics of the neuronal submembrane skeleton, and the morphology and function of neurons. Thus, we next investigated the neuronal effects of human βII-spectrin mutations using a structure-function rescue approach in βII-SpKO cortical neurons.

First, we expressed WT and mutant GFP-βIISp together with mCherry in day in vitro (DIV) 3 βII-SpKO cortical neurons^26^ and evaluated their neuronal growth at DIV8. We also evaluated mCherry-expressing WT *(Sptbn1^flox/flox^* /+) and heterozygous *(Sptbn1^flox/+^;*Nestin-Cre; henceforth abbreviated as βII-SpHet) neurons grown in parallel. As previously observed^26^, neuronal growth, quantified through axonal length, was severely impaired in βII-SpKO neurons, but restored upon expression of WT GFP-βIISp (Fig. 5a,b and Extended Data Fig. 4a). βII-SpHet neurons grew to roughly only half the length of WT neurons, but their axons were at least twice as long as βII-SpKO neurons (Fig. 5a,b). Most of the βII-spectrin mutants failed to rescue axonal length except for p.G1398S and p.E1886Q, which restored growth to WT levels, while p.A1086T and p.E1110D restored length to heterozygous levels (Fig. 5a,b and Extended Data Fig. 4a). Remarkably, some of the aberrant morphological features observed in HEK293 cells were also present and often more markedly displayed in neurons expressing mutant GFP-βIISp. As shown above, p.C183*, p.Y190_R216del, p.G205S, and p.G205D GFP-βIISp mutants were almost exclusively distributed in large protein aggregates localized to the neuronal cell bodies and in some processes (Fig. 3b and Extended Data Fig. 5a). All other mutants within the CH domain invariably produced extensive aberrant membrane features in the form of lamellipodia and filopodia around the cell body and along the neuronal processes (Fig. 6a and Extended Data Fig. 5a). Similarly, the p.A850G mutant resulted in cell bodies and neuronal processes with expanded membranes extensively decorated with filopodia-like protrusions, while the p.R411W mutant led to a milder phenotype (Extended Data Fig. 5a). Neuronal membrane expansion was accompanied by a shift in the boundaries of actin and αII-spectrin distribution (Fig. 6a). These results confirm that clinically relevant βII-spectrin mutations can cause marked disruptions in cell morphology, likely driven by disruptions in the submembrane cytoskeleton organization and dynamics, which may be a pathogenic factor in *SPTBN1* syndrome.

**Fig. 5:**
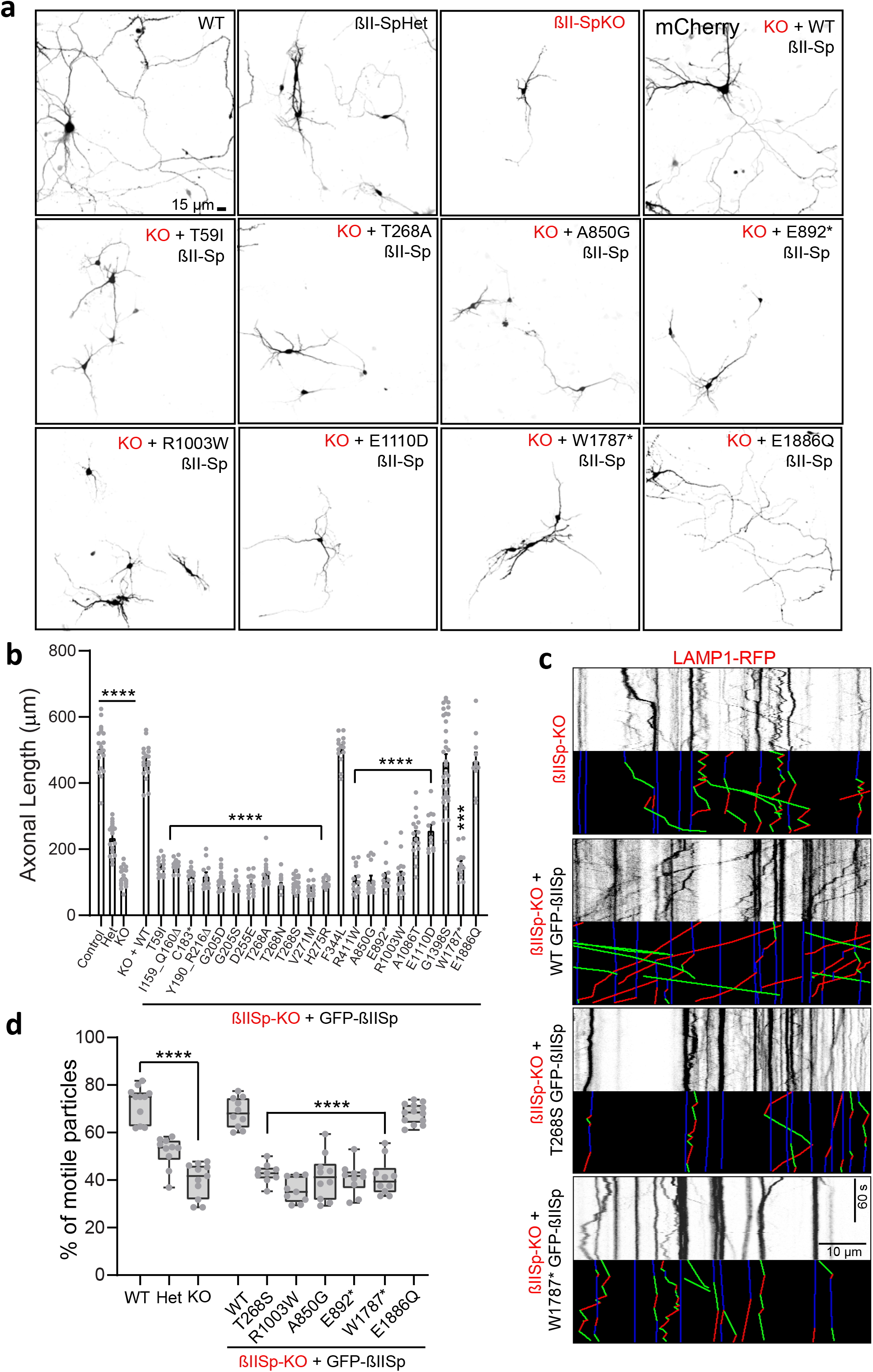
*SPTBN1* variants affect neuronal axonal growth and organelle transport. **a**, Images of DIV8 βII-SpWT, βII-SpHet, and βII-SpKO neurons transfected at DIV3 with mCherry. A subset of βII-SpKO neurons was co-transfected with GFP-βIISp and mCherry plasmids. Scale bar, 15 μm. **b**, Axonal length of βII-SpWT, βII-SpHet, βII-SpKO, and rescued βII-SpKO DIV8 neurons (n=12-34 neurons/genotype) compiled from three independent experiments. Data represent mean ±SEM. One-way ANOVA with Dunnett’s post hoc analysis test for multiple comparisons, ****p < 0.0001. **c**, Kymographs showing the mobility of RFβ-tagged LAMP1-positive cargo in axons from DIV8 βII-SpKO and rescued βII-SpKO neurons. Analyzed trajectories are shown with a color code with green for anterograde, red for retrograde, and blue for static vesicles. Scale bar, 10 μm and 60 s. **d**, Quantification of percent of motile LAMP1-RFβ-positive cargo in axons from βII-SpWT, βII-SpHet, βII-SpKO, and rescued βII-SpKO neurons. The box and whisker plots represent data from minimum to maximum collected in n=9-13 axons per genotype. One-way ANOVA with Dunnett’s post hoc analysis test for multiple comparisons, ****p < 0.0001.

**Fig. 6:**
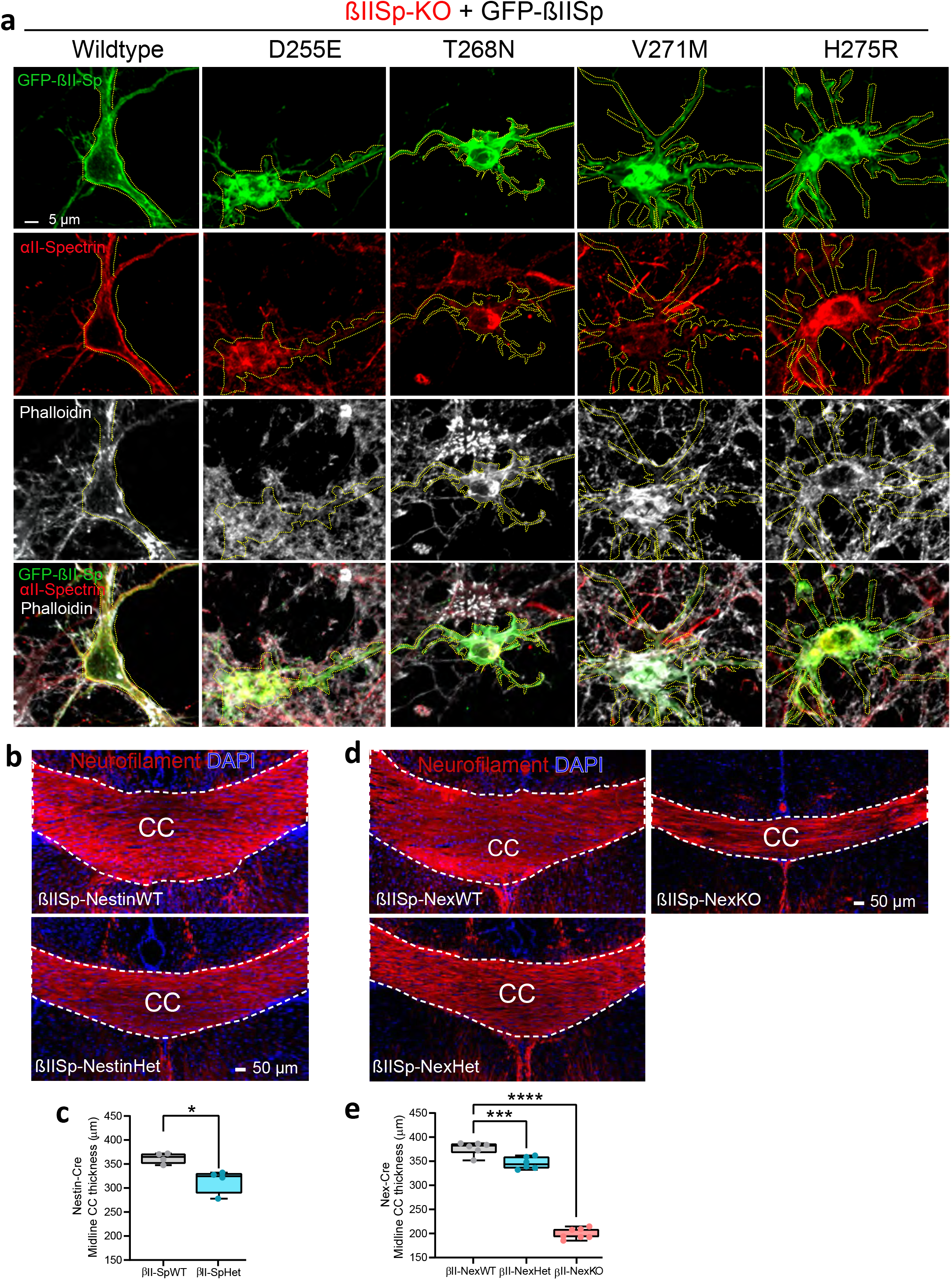
βII-spectrin deficiency disrupt proper neuronal development in cortical neuronal cultures and mouse brains. **a**, Images of DIV8 βII-SpKO cortical neurons rescued with WT GFP-βIISp or with GFP-βIISp bearing mutations within the distal portion of the CH2 domain. Neurons were stained for actin (phalloidin) and endogenous αII-spectrin. Yellow dotted lines demark the cell edge. Scale bar, 5 μm. **b**, Images of PND25 βII-SpWT and βII-SpHet brains stained for neurofilament to label axons and DAPI. Scale bar, 50 μm. **c**, Quantification of CC thickness at the brain midline. (n=4 mice/genotype). Data represent mean ± SEM. Unpaired t-test, *p < 0.05. **d**, Images of PND25 βII-SpNexWT, βII-SpNexHet and βII-SpNexKO brains stained for neurofilament to label axons and DAPI. Scale bar, 50 μm. White dotted lines in b and d denote the position and boundaries of the corpus callosum (CC). **e**, Quantification of CC thickness at the brain midline. (n=6-7 mice/genotype). Data in c and e represent mean ± SEM. One-way ANOVA with Dunnett’s post hoc analysis test for multiple comparisons, ***p < 0.001, ****p < 0.0001.

Organelle transport is essential for the maintenance of neuronal processes and viability of neurons and defects in transport can contribute to the pathology of several neurological diseases^45^. We previously showed that βII-spectrin promotes normal organelle axonal transport independently of its role assembling the MPS^26^. Expression of WT βII-spectrin in cultured βII-spectrin null cortical neurons rescues the processivity, motility, and flux of synaptic vesicles and lysosomes^26^. To evaluate the effects of selected βII-spectrin mutations on axonal transport, we tracked the dynamics of red fluorescent protein (RFP)-tagged LAMP1 (an endosome/lysosome vesicles marker) in control, βII-SpKO, and βII-SpHet cortical neurons using time-lapse video microscopy. As previously observed^26^, loss of βII-spectrin impairs the bidirectional motility of LAMP1-RFβ-positive vesicles and causes significant deficits in their run length and retrograde velocity (Fig. 5c,d and Extended Data Fig. 4b,c). Remarkably, βII-spectrin haploinsufficiency causes similar deficits (Fig. 5c,d and Extended Data Fig. 4b,c), indicating that 50% reduction of βII-spectrin levels is not sufficient to maintain normal organelle transport. As expected^26^, deficits in transport of lysosomes in βII-SpKO neurons are rescued by expression of WT GFPβIISp. However, selected mutants that do not rescue axonal length also fail to restore normal organelle dynamics (Fig. 5c,d and Extended Data Fig. 4b,c). Within the mutants tested, p.E892* and p.W1787* lack the PH domain, which is required for βII-spectrin coupling to organelle membranes and normal organelle transport^25^. It is possible that the abnormal binding to molecular partners observed in other mutants unable to rescue organelle dynamics interfere with the formation of complexes between βII-spectrin and molecular motors, its coupling to organelle membranes, or its cytosol to MPS partitioning.

Collectively, our results suggest that human βII-spectrin mutations we report likely cause *SPTBN1* syndrome through molecular and cellular mechanisms that include the individual or combined effects of toxic protein aggregation, disruption of intracellular organelle transport, insufficient axonal growth, and aberrant cytoskeletal organization and dynamics, which in turn may affect neuronal connectivity and function.

### *SPTBN1* variant classification

The *SPTBN1* variants described in this study were classified using the 2015 ACMG Guidelines^46^ and interpretation recommendations^47-49^ and are listed in Supplementary Table 2 with a summary of functional evidence herein. Of the 28 unique variants in the cohort, 14 were classified as pathogenic, 12 as likely pathogenic, and two as variants of uncertain significance (VUS). Importantly, proband P10 has two *de novo* variants in *cis* in *SPTBN1*, p.T268A and p.F344L. The p.T268A variant has two allelic variants p.T268N and p.T268S and functional studies suggesting a damaging effect support a pathogenic classification. The p.F344L variant is classified as a VUS since it is in *cis* with a pathogenic variant and showed no significant differences from wild type in functional studies, thus its contribution to the phenotype of this individual is unclear.

### βII-spectrin haploinsufficiency causes cell-autonomous deficits in neuronal connectivity

βII-spectrin is widely expressed in both neurons and in brain non-neuronal cells^50^. βII-spectrin loss in both neurons and glial cells of βII-SpKO mice results in significant reduction of long-range axonal tracts in the cerebellum and of those tracts connecting cerebral hemispheres, including the CC^26^. These white matter connectivity deficits are likely caused by the impaired axonal growth of neurons lacking βII-spectrin^26^. Since βII-spectrin haploinsufficiency affects axonal growth *in vitro* (Fig. 5a, b), we next assessed cortical axonal connectivity in βII-SpHet mice. Consistent with a diminished axonal growth, PND25 βII-SpHet mice exhibit callosal hypoplasia (Fig. 6b,c). CC thinning is also detected by MRI in three of the probands in this cohort (P2, P10, and P28) (Fig. 1d and Table 1 and Supplementary Note), which further implicates βII-spectrin in regulating brain cytoarchitecture. Deficits in connectivity of long axonal tracts can also result from defects in neuronal migration and axonal pathfinding, which in turn can be affected by non-neuronal cells^51^. To determine the neuron-specific effects of βII-spectrin depletion on cortical wiring, we generated mice lacking βII-spectrin only in cortical and hippocampus projection neurons by crossing *Sptbn1^fhx/flox^* to Nex-Cre^52^ animals *(Sptbn1^flox/flox^*;Nex-Cre; henceforth referred to as βIISp-Nex KO) (Extended Data Fig. 5b). βII-spectrin loss or haploinsufficiency only in projection neurons is sufficient to induce CC hypoplasia (Fig. 6c,d). These results suggest that partial βII-spectrin LOF can produce neuronal miswiring in the cortex and those defects are at least in part neuron-autonomous.

### βII-spectrin deficiency causes developmental and behavioral deficits in mice

Individuals bearing *SPTBN1* variants exhibit a wide range of facial dysmorphisms, brain growth defects, including microcephaly and macrocephaly, and DD (Table 1, Supplementary Note). We found that embryonic day 19 (E19) βII-SpKO mice have enlarged head circumference, and both E19 βII-SpKO and βII-SpHet animals exhibit increased distance between the eyes (Fig. 7a-c), consistent with the observed hypertelorism in some of the patients (Fig. 1c and Supplementary Note). In line with reported DD in patients, βII-SpKO mice show arrested development (Fig. 7d,e)^26^. in addition, βII-spectrin haploinsufficiency is sufficient to yield animals of an intermediate body size and weight (Fig. 7d-f). The global DD changes observed in mice with βII-spectrin deficits arise in part due to neuronal-autonomous effects, given that they are also observed in βIISp-NexKO mice that only lack the protein in cortical and hippocampal projection neurons (Extended Data Fig. 6a).

**Fig. 7:**
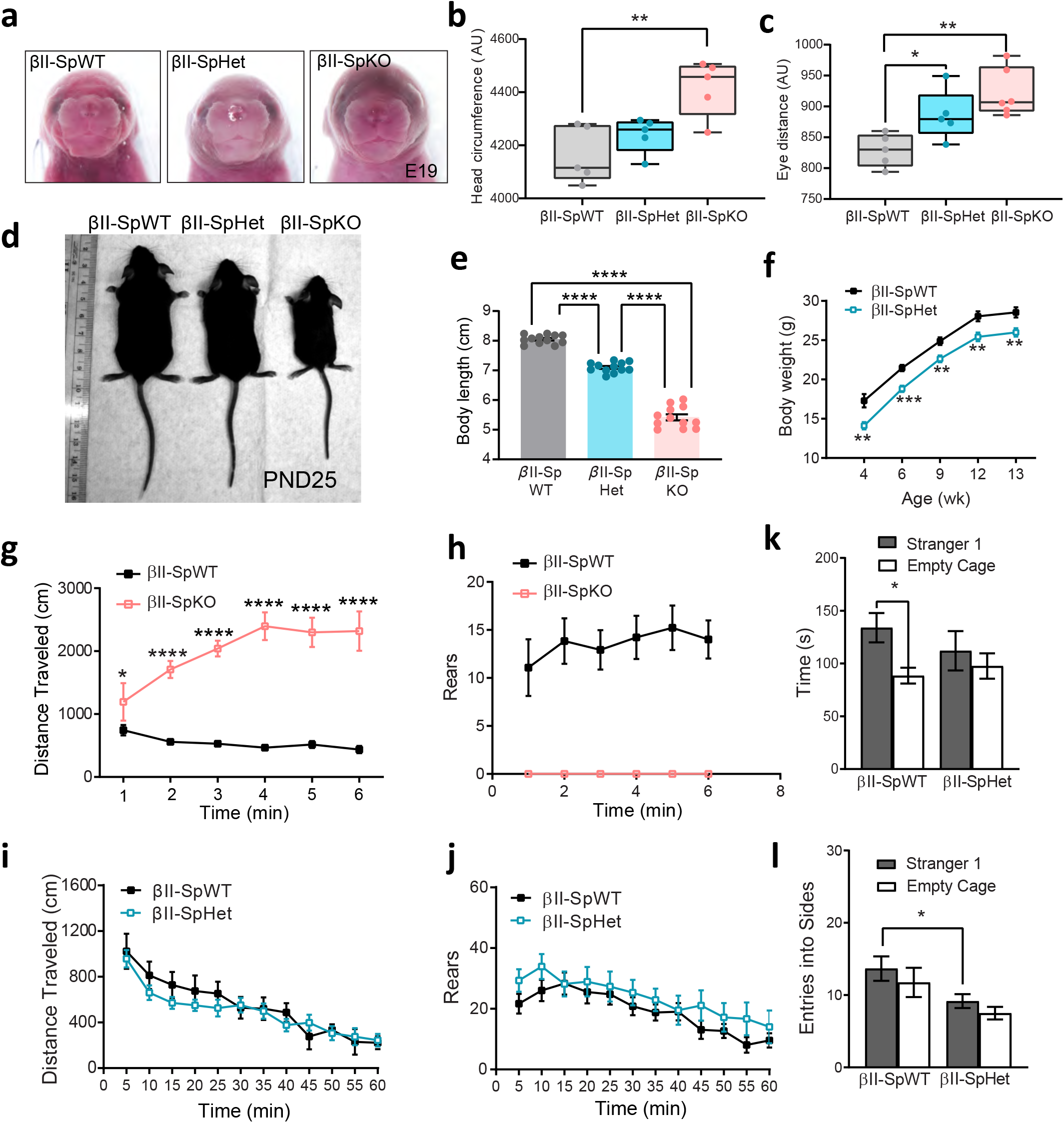
βII-spectrin deficiency causes developmental and behavioral deficits in mice. **a**, Images of male E19 βII-SpWT, βII-SpHet, and βII-SpKO embryos. **b, c**, Quantification of head circumference (b) and eye distance (c) at E19. Data represents mean ± SEM (n=5-6 embryos/genotype). One-way ANOVA with Dunnett’s post hoc analysis test for multiple comparisons, *p < 0.05, ** p< 0.01. **d**, Images of male PND25 wildtype (βII-SpWT) mice and mice with partial (βII-SpHet) and complete (βII-SpKO) loss of βII-spectrin in neural progenitors. **e**, Body length at PND25 for indicated genotypes. Data represent mean ± SEM (n=12 male mice/genotype). One-way ANOVA with Dunnett’s post hoc analysis test for multiple comparisons, ****p <0.0001. **f**, Growth curve (as body weight) of βII-SpWT and βII-SpHet mice. Data represent mean ± SEM (n=12 male mice/genotype). Unpaired t-test, ** p< 0.01, ***p < 0.001. **g**, Locomotor activity and **h**, rearing assessed during a 30-minutes open-field test in PND30 βII-SpWT and βII-SpKO mic. Data in g and h represent mean ± SEM (n=15 βII-SpWT and n=5 βII-SpKO male mice). Unpaired t-test, * p< 0.05, ****p <0.0001. Statistical comparisons were not conducted for h due to zero scores in the βII-SpKO group. **i, j**, Locomotor activity measured as distance traveled (i) and number of rears (j) during a one-hour test in a novel open field. **k**, Lack of social preference in βII-SpHet mice during a three-chamber choice task. **l**, Decreased entries by ßII-SpHet mice into a chamber with stranger mouse. Data represent mean ± SEM (n=12 male mice/genotype). Unpaired f-test, *p< 0.05.

Since individuals carrying *SPTBN1* variants present with various behavioral phenotypes, including ASD, ADHD as well as learning and mild motor deficits (Table 1, Supplementary Note), we assessed behavioral effects of brain βII-spectrin deficiency in mice. First, we evaluated the effects of complete LOF using βII-SpKO mice. Because these animals do not survive longer than five weeks^26^, they were only challenged with open field and acoustic startle tests at PND30. βII-SpKO mice had overt hyperactivity at every time point during the open field test (Fig. 7g) and profound deficits in rearing, a response requiring good hind limb function and balance (Fig. 7h). βII-SpKO mice also showed decreases in startle response amplitudes in the acoustic startle test, but normal levels of prepulse inhibition (PPI) (Extended Data Fig. 6b,c), suggesting that reduced startle responses were due to motor deficits, rather than alterations in auditory function or sensorimotor gating. This is consistent with impaired motor abilities likely due to the severe loss of cerebellar connectivity in these mice^26^.

Our clinical, cellular, and animal data indicates that βII-spectrin haploinsufficiency, or the altered function of only one copy of *SPTBN1* due to GOF or dominant-negative effects, is sufficient to cause a neurodevelopmental disorder. Thus, we next characterized behavioral phenotypes of βII-SpHet mice, whose normal lifespan allowed for an expanded battery of tests. in contrast to βII-SpKO mice, βII-SpHet animals had normal activity during an open field test (Fig. 7i,l). Further, βII-SpHet and control mice had comparable performance in an acoustic startle test for PPI and in the rotarod test (Extended Data Fig. 6d-f), indicating that expression of half levels of βII-spectrin is sufficient to rescue motor problems. βII-SpHet mice also exhibited normal spatial and reversal learning in the Morris water maze test (Extended Data Fig. 6g,h). On the other hand, in the 3-chamber choice test, βII-SpHet demonstrated no preference for spending more time in proximity to a stranger mouse versus an empty cage and made significantly fewer entries into the side containing the stranger mouse (Fig. 7k,l). These genotype differences were not observed in the subsequent test for social novelty preference, in which βII-SpHet and βII-SpWT littermates demonstrated the typical shift in preference to the newly introduced stranger 2 (Extended Data Fig. 6k,l). Notably, there was a non-significant trend for the βII-SpHet mice to make fewer entries than the littermate controls in the social novelty test. The lack of sociability in the βII-SpHet mice was not associated with changes in anxiety-like behavior or olfactory function (Extended Data Fig. 7m). Overall, these results suggest that βII-spectrin LOF impairs global development and has a selective impact on social motivation and reward that may contribute to the autistic features and social behavior impairments manifested in some affected individuals.

## Discussion

In this study, we report for the first time the identification of *de novo SPTBN1* variants in individuals as a cause of a neurodevelopmental disorder most commonly characterized by motor and speech delays, ID, and various neurologic and behavioral comorbidities. In addition to DD and ID, eleven individuals in our cohort have been diagnosed with ADD/ADHD and six with ASD, with three having co-occurrence. This observation is consistent with a recent WES study of a Danish cohort of approximately 8,000 children with ASD and/or ADHD and 5,000 controls that identified *SPTBN1* as a top hit among genes with rare truncating variants co-occurring in these disorders at a significantly higher rate than in controls^53^. *SPTBN1* variants had previously been reported in probands with ASD^29^, Tourette^28^, and DD (all included in our study). Noteworthy, βII-spectrin’s canonical partner ankyrin-B is encoded by high confidence ASD gene *ANK2^27^* and some ASD patients with *ANK2* variants also exhibit ID^54^. Loss of ankyrin-B isoforms in mice result in axonal transport deficits^55^ and developmentally regulated defects in brain connectivity^54,55^, two overlapping phenotypes we observed in our βII-spectrin mouse models. Although ankyrin-B and βII-spectrin are independent modulators of axonal transport^26^, *SPTBN1* and *ANK2* may otherwise converge through mechanisms that affect other neuronal functions. For example, loss of ankyrin-B affects the polarized distribution of βII-spectrin in neurites, which gives rise to its more even portioning between axons and dendrites causing a higher than normal prevalence of the MPS in dendrites^56^. Conversely, disruption of the MPS due to loss of βII-spectrin^24,26^ may disrupt the periodic distribution of ankyrin-B and its membrane partners in axons^54^, which together may be essential for critical signal transduction events^57^. In addition to their strong correlation with DD, our results together with these observations support the association of *SPTNB1* pathogenic variants with ASD and ADHD.

Seizures and epilepsy were other noticeable re-occurring phenotypes in our cohort. That *SPTBN1* variants may have epileptogenic effects is not surprising, given the strong association of *de novo* and inherited variants in the partner gene *SPTAN1* (αII-spectrin) with epileptic syndromes^5,16-21^. Although the precise pathogenic mechanism of *SPTAN1* in epilepsy is unknown, αII-spectrin protein aggregation has been reported for several of the putative pathogenic variants^16,20^. As we show above, αII-spectrin cellular distribution can be disrupted by mutant βII-spectrin to cause these partners to co-aggregate, or otherwise continue to associate in aberrant cellular patterns. Since βII- and αII-spectrin are critically involved in localizing and stabilizing ion channels^1-3^, going forward it will be critical to elucidate whether these tightly intertwined partners share pathways disrupted in channelopathies underlying seizures and epilepsy.

Besides the widely shared DD phenotype in our cohort, further supporting evidence of the pathogenicity of *SPTBN1* variants is the re-occurrence of *de novo* variants in the same amino acid position in unrelated individuals which are not found in the general population. These individuals share other co-occurring clinical manifestations, but also diverge in some of the clinical presentations, which may originate in part by differences in the identity of the amino acid substitution, sex, age, and genetic background. Another striking indicator of convergence in the pathogenic mechanism of the βII-spectrin mutations we report is their partial clustering (14 of 28) within the CH domains. The region of *SPTBN1* encoding the CH domains has a higher degree of missense variant constraint in the population (ExAC v.10)^58^, indicating the importance of the CH domains for protein function and supporting the pathogenicity of the variants within. Our cellular and biochemical findings suggest that CH domain mutants generally affect βII-spectrin’s interaction with F-actin and αII-spectrin and result in modified spectrin/actin cytoskeleton dynamics and cellular morphology. The aberrant accumulation of mutant βII-spectrin within cytosolic aggregates suggests that a subset of the CH mutations introduce destabilizing effects on the protein structure, which is supported by our structural modeling. These changes in βII-spectrin distribution, as well as in binding to submembrane cytoskeleton partners, likely underlie GOF effects, such as aberrant neuronal membrane morphology, and contribute to LOF deficits, such as impaired organelle transport and reduced axonal growth. In turn, these cellular defects likely result in the deficient or aberrant brain connectivity and function observed in βII-spectrin-deficient mice and in patients. Interestingly, pathogenic CH domain variants have been reported in βI-spectrin^59^, which cause spherocytosis, and PIII-spectrin^4,13^, which leads to cerebellar ataxia, DD, and ID, and have been shown to affect F-actin binding^41^. Together with our results, this evidence indicates that the abnormal modulation of actin binding by CH domain variants likely constitute a conserved pathogenic mechanism in spectrinopathies.

Like in other spectrinopathies^4-23^, missense mutations affecting SR are likely to be disease-causing in the *SPTBN1* syndrome, although the molecular mechanisms are not fully understood. For example, it is not clear how p.A850G phenocopies the cellular phenotypes caused by some of the CH domain mutants. It is possible that this mutant affects βII-spectrin/F-actin dynamics through allosteric mechanisms or dominant negative effects due to overexpression. Alternatively, this and the other SR mutants may disrupt βII-spectrin association with undefined binding partners or its coupling to organelles and motor proteins, which may explain their detrimental effect on axonal growth^26^. Our cellular assays failed to identify a potential pathogenic mechanism for a small subset of *SPTBN1* variants. However, it is possible that these variants affect other less explored neuronal βII-spectrin roles, such as dendritic and postsynaptic development and function, which are associated with ASD^29^. Additionally, given the wide expression of βII-spectrin in non-neuronal brain cells, it will be of interest to assess if their function is affected by *SPTBN1* variants. It is likely that the clinical variability is at least partly rooted in the multifunctionality and ubiquitous expression of βII-spectrin, although we cannot rule out that some clinical manifestations unique to affected individuals in the cohort may be caused by an alternate etiology. For example, a few individuals in the cohort have additional genetic variants that might be contributing to their clinical phenotype. Proband P19 has a pathogenic *NF1* variant (NM_000267.3:c.3449C>G; p.S1150*) and has neurofibromatosis, which could also have associated learning disabilities, but likely would not explain the behavioral challenges and autism seen in this individual. Proband P27 has a variant in *GNB1* (NM_001282539.1:c.700-1G>T) inherited from her mother also affected with delays. However, the *SPTBN1* variant was not present in the mother, and could be *de novo* or paternal as her father has moderate ID, suggesting both variants could be contributory. Finally, given the critical roles βII-spectrin plays in other organs^60,61^ and its association with other non-neurological disorders, including clinical presentations beyond the nervous system in patients in our cohort, the *SPTBN1* syndrome warrants thorough clinical assessment and further studies beyond the brain.

## Materials and methods

### Identification of Pathogenic *SPTBN1* Variants

Pathogenic variants in *SPTBN1* were identified by whole exome or genome sequencing performed on whole blood DNA from probands identified through diagnostic clinical practice or Institutional Review Board approved research studies. Affected individuals were identified through professional communication, connections through GeneMatcher^62^, and by searching the Undiagnosed Diseases Network (UDN) and the Deciphering Developmental Disorders (DDD) Research Study^29^ repositories. Variants were reported according to standardized nomenclature defined by the reference human genome GRCh37 (hg19) and *SPTBN1* transcript GenBank: NM_003128.2. The minor-allele frequency of each variant was determined from genomic sequencing data derived from the gnomAD.

### Patient consent

Patient consent for participation and phenotyping was obtained through the referring clinical teams. Referring clinicians were requested to complete a comprehensive questionnaire that was based upon our current understanding of the phenotypic associations of *SPTBN1*. They included sections related to neurodevelopmental screening, behavior, dysmorphology, muscular, cardiac, and other systemic phenotypic features. Consent and collection of information conformed to the recognized standards of the Declaration of Helsinki and approved by local Institutional Review Boards.

### Variant interpretation and classification

*SPTBN1* variants were interpreted using the NM_003128.2 transcript and splice variants were evaluated using SpliceAI^27^ to predict the most likely mRNA splicing outcome. The *SPTBN1* variants identified in this study were classified according to the ACMG 2015 Guidelines^46^. Based on the recommendations of PVS1 loss-of-function criterion under the ACMG/AMP specifications^47^, PVS1_strong was used as a maximum weight of evidence. This is appropriate for this criterion as we have shown moderate clinical validity^48^, unrelated probands with a consistent phenotype, and robust functional evidence showing that these nonsense variants remove downstream portions of the protein known to be essential for protein function, and that both null and haploinsufficient mouse models recapitulate disease phenotypes. The maximum weight of functional evidence (PS3) used was moderate under the ACMG/ACMP specifications^49^.

### Mouse lines and animal care

Experiments were performed in accordance with the guidelines for animal care of the Institutional Animal Care and Use Committee of the University of North Carolina at Chapel Hill. To generate neural progenitor-specific βII-spectrin null *(Sptbn1^flox/flox^*/Nestin-Cre, βIISp-KO) and haploinsufficient *(Sptbn1^flox/^+/Nestin-Cre*, βIISp-Het) mice, *Sptbn1^flox/fhx^* animals, a gift from Dr. Mathew Rasband^50^, were crossed with the Nestin-Cre mouse line [B6.Cg-Tg(Nes-cre)1Kln/J, stock number 003771; The Jackson Laboratory]. *Sptbn1^flox/fhx^* animals negative for the Cre transgene were used as littermate controls in all experiments. Mice lacking βII-spectrin in cortical projection neurons *(Sptbn1^flox/flox^/Nex-Cre*, βIISp-Nex KO) were generated by crossing *Sptbn1^flox/flox^* and Nex-Cre, a gift from Dr. Klaus-Armin Nave^52^, animals for multiple generations. All mice were housed at 22°C ± 2°C on a 12-hour-light/12-hour-dark cycle and fed ad libitum regular chow and water.

### Generation of human βII-spectrin mutations

The human βII-spectrin cDNA was subcloned into peGFP-C3 vector (Clontech) using HindIII and SacI sites to generate the peGFP-βIISp plasmid. For purification of full-length βII-spectrin proteins, both a prescission protease site (LEVLFQGP) and a 6x histidine tag were respectively introduced between the GFP and start codon and before the C-terminal stop codon of peGFP-βII-spectrin using site-directed mutagenesis to generate the peGFP-PP-βII-Sp-6xHis construct. peGFP-βIISp and peGFP-PP-βII-Sp-6xHis plasmids bearing the human mutations included in the study were generated using the In-Fusion HD Cloning Plus system (Takara) and primers specific for each mutation site (Supplementary Table 3). All plasmids were verified by full-length sequencing.

### Plasmids

Plasmid used in transfection experiments included: pLAMP1-RFP (Addgene plasmid #1817, gift from Walther Mothes), pmCherry-C1 (Clontech) and peGFP-C3 vector (Clontech). To generate mCherry-tagged αII-spectrin (pmCherry-αIISp), the cDNA sequence of human αII-spectrin (NM_001130438.3) was amplified by PCR as a BsrGI/XhoI fragment and cloned into the corresponding sites of pmCherry-C1 (Clontech). peGFP-C3-Y1874A-βII-spectrin and HA-tagged 220 kDa ankyrin-B (pAnkB-3X HA) plasmids were previously reported.^26^ All plasmids were verified by full-length sequencing prior to transfection.

### Antibodies

Affinity-purified rabbit antibodies against GFP and βII-spectrin, used at a 1:500 dilution for immunohistochemistry and 1:5000 for western blot, were generated by Dr. Vann Bennett laboratory and have been previously described.^26,51^ Other antibodies used for western blot analysis and immunoprécipitation included mouse anti-GFP (1:1000, #66002-1-Ig,), rabbit anti-GFP (1:1000, #50430-2-AP), rabbit anti-HA tag (1:1000, #51064-2-AP), and mouse anti-6*His tag (1:1000, # 66005-1-Ig) all from Proteintech, and rabbit anti-mCherry (1:2000, #ab24345) from Abcam. Commercial antibodies used for immunofluorescence included mouse anti-neurofilament (1:200, # 837801) from BioLegend and chicken anti-GFP (1:1000, #GFP-1020) from Aves. Secondary antibodies purchased from Life Technologies were used at 1:400 dilution for fluorescence-based detection by confocal microscopy, and included donkey anti-rabbit IgG conjugated to Alexa Fluor 568 (#A10042), donkey anti-mouse IgG conjugated to Alexa Fluor 488 (#A21202), goat anti-chicken conjugated to Alexa Fluor 488 (#A11039), and donkey anti-rat IgG conjugated to Alexa Fluor 647 (#A21247). Fluorescent signals in western blot analysis were detected using goat anti-rabbit 800CW (1:15000, #926-32211) and goat anti-mouse 680RD (1:15000, #926-68070) from LiCOR.

### Neuronal culture

Primary cortical neuronal cultures were established from E17 mice. Cortices were dissected in Hibernate E (Life Technologies) and digested with 0.25% trypsin in HBSS (Life Technologies) for 20 min at 37°C. Tissue was washed three times with HBSS and dissociated in DMEM (Life Technologies) supplemented with 5% fetal bovine serum (FBS, Genesee), and gently triturated through a glass pipette with a fire-polished tip. Dissociated cells were filtered through a 70 μm cell strainer to remove any residual non-dissociated tissue and plated onto poly-D-lysine-coated 1.5 mm coverglasses or dishes (MatTek) for transfection and time-lapse microscopy imaging. For all cultures, media was replaced 3 hours after plating with serum-free Neurobasal-A medium containing B27 supplement (Life Technologies), 2 mM Glutamax (Life Technologies), and penicillin/streptomycin (Life Technologies). 5 μM cytosine-D-arabinofuranoside (Sigma) was added to the culture medium to inhibit the growth of glial cells three days after plating. Neurons were fed twice a week with freshly made culture medium until use.

### Plasmid transfection for time-lapse live imaging and immunofluorescence analysis

For time-lapse imaging experiments DIV5 cortical neurons were co-transfected with 1μg of each pLAMP1-RFP and peGFP-βIISp plasmids using lipofectamine 2000 (Life Technologies) and imaged 48-96 hours after transfection. For experiments that evaluate axonal length, DIV3 control and βIISp-Het neurons were transfected with 500 ng of pmCherry-C1 and 1μg of peGFP-C3. βII-SpKO neurons were transfected with 500 ng of pmCherry-C1 and 1μg of peGFP-βIISp recue plasmids bearing full-length wildtype of mutant β2-spectrin. Neurons were processed for immunofluorescence 5 days after transfection. Immunofluorescence evaluations of βII-spectrin distribution in HEK293 cells was conducted in cells transfected with l00 ng of peGFP-βIISp plasmids, or co-transfected with 100 ng of each peGFP-βIISp and pmCherry-αIISp plasmids 48 hours post-transfection.

### Plasmid transfection for biochemistry analysis

All transfections were conducted in HEK293 cells grown in 10 cm culture plates using the calcium phosphate transfection kit (Takara). To purify full-length βII-spectrin proteins, cells were transfected with 8 μg of peGFP-PP-βII-Sp-6xHis plasmids. To determine levels and stability of Rll-spectrin proteins, HEK293T cells were co-transfected with 8 μg of eGFP-PP-βII-Sp-6xHis and 4 μg of pmCherry-C1 plasmids. To determine interaction between ankyrin-B and βII-spectrin, cells were co-transfected with 8μg of each peGFP-PP-βII-Sp-6xHis and pAnkB-3X HA plasmids. For assessment of binding between βII-spectrin and αII-spectrin, cells were separately transfected with 8 μg of peGFP-PP-βII-Sp-6xHis or 4 μg peGFP-C3 and 8 μg of pmCherry-αIISp.

### Histology and immunohistochemistry

Brains from mice two-weeks and older were fixed by transcardial perfusion with phosphate-buffered saline (PBS) and 4% paraformaldehyde (PFA) followed by overnight immersion in the same fixative. Brains from PND0-PND14 mice were fixed by direct immersion in 4% PFA for 36 hours. After fixation, brains were rinsed with PBS, transferred to 70% ethanol for at least 24 hours, and paraffin-embedded. 7-μm coronal and sagittal brain sections were cut using a Leica RM2155 microtome and mounted on glass slides. Sections were analyzed by hematoxylin and eosin (H&E) staining or immunostaining. For antibody staining, sections were deparaffinized and rehydrated using a standard protocol of washes: 3 x 3-min Xylene washes, 3 x 2-min 100% ethanol washes, and 1 x 2-min 95%, 80%, and 70% ethanol washes followed by at least 5 min in PBS. Sections were then processed for antigen retrieval using 10 mM sodium citrate, pH 6 in the microwave for 20 min. Sections were allowed to cool, washed in PBS, and blocked using antibody buffer (2% bovine serum albumin (BSA), 1% fish oil gelatin, 5% donkey serum, and 0.02% Tween 20 in PBS) for 1 hour at room temperature. Tissue sections were then subsequently incubated overnight with primary antibodies at 4°C and with secondary antisera for 1.5 hours at 4°C, washed with PBS, and mounted with Prolong Gold Antifade reagent (Life Technologies). Neuronal cultures and HEK293 cells were washed with cold PBS, fixed with 4% PFA for 15 min, and permeabilized with 0.2% Triton-X100 in PBS for 10 min at room temperature. Neurons and HEK293 cells were blocked in antibody buffer for 1 hour at room temperature and processed for fluorescent staining as tissue sections. For actin labeling, Alexa Fluor 568- or Alexa Fluor 633-conjugated phalloidin (1:100) was added to the secondary antibody mix. DAPI was added to the last PBS rinse for nuclei staining.

### Immunoblots

Protein homogenates from mouse brains or transfected cells were prepared in 1:9 (wt/vol) ratio of homogenization buffer (8M urea, 5% SDS (wt/vol), 50mM Tris pH 7.4, 5mM EDTA, 5mM N-ethylmeimide, protease and phosphatase inhibitors) and heated at 65°C for 15 min to produce a clear homogenate. Total protein lysates were mixed at a 1:1 ratio with 5x PAGE buffer (5% SDS (wt/vol), 25% sucrose (wt/vol), 50mM Tris pH 8, 5mM EDTA, bromophenol blue) and heated for 15 min at 65°C. Samples were resolved by SDS-PAGE on 3.5-17.5% acrylamide gradient gels in Fairbanks Running Buffer (40mM Tris pH 7.4, 20mM NaAc, 2mM EDTA, 0.2%SDS (wt/vol)). Proteins were transferred overnight onto 0.45 μm nitrocellulose membranes (#1620115, BioRad) at 4°C. Transfer efficiency was determined by Ponceau-S stain. Membranes were blocked in TBS containing 5% non-fat milk for 1 hour at room temperature and incubated overnight with primary antibodies diluted in antibody buffer (TBS, 5% BSA, 0.1% Tween-20). After 3 washes in TBST (TBS, 0.1% Tween-20), membranes were incubated with secondary antibodies diluted in antibody buffer for two hours at room temperature. Membranes were washed 3x for 10 minutes with TBST and 2x for 5 minutes in TBS. Protein-antibody complexes were detected using the Odyssey® CLx Imaging system (LI-COR).

### Immunoprecipitation

For immunoprecipitation experiments, total protein homogenates from transfected HEK293 cells were prepared in TBS containing 150 mM NaCl, 0.32 M sucrose, 2 mM EDTA, 1% Triton X-100, 0.5% NP40, 0.1% SDS, and compete protease inhibitor cocktail (Sigma). Cell lysates were incubated with rotation for 1 hour at 4°C and centrifuged at 100,000 x g for 30 min. Soluble fractions were collected and precleared by incubation with Protein-G magnetic beads (#1614023, Bio-Rad) for 1 hour in the cold. Samples were subjected to immunoprecipitation in the presence of protein-G magnetic beads/antibody or protein-G magnetic beads/isotype control complexes overnight at 4°C. Immunoprecipitation samples were resolved by SDS-PAGE and western blot and signal detected using the Odyssey® CLx imaging system.

### Purification of full-length βII-spectrin proteins

Ten 10-cm plates of HEK293 cells expressing each peGFP-PP-βII-Sp-6xHis construct were used per purification. Total protein homogenates from transfected HEK293 cells were prepared in TBS containing 150 mM NaCl, 0.32 M sucrose, 2 mM EDTA, 1% Triton X-100, 0.5% NP40, 0.1% SDS, and complete protease inhibitor cocktail (Sigma) (IP buffer). Cell lysates were incubated with rotation for 1 hour at 4°C and centrifuged at 100,000 x g for 30 min. Soluble fractions were incubated overnight with Protein A/G magnetic beads (#88802, Life Technologies) coupled to GFP antibodies with rotation at 4°C. Beads were extensively washed with IP buffer, followed by washes in TBS containing 300 mM NaCl, and TBS. Full-length βII-spectrin proteins were eluted from GFP-protein A/G magnetic beads by incubation with HRV-3C protease, which cleaves between GFP and the start codon of βII-spectrin in prescission protease buffer (25 mM HEPES, 150 mM NaCl, 1 mM EDTA, 1 mM DTT) for 36 hours at 4°C. The efficiency of cleavage and purity of the eluates was analyzed by western blot using validated antibodies specific for βII-spectrin and GFP and 6*His tags, and by Coomassie blue stain. Eluates were concentrated using Pierce™ Protein Concentrators PES.

### Pulldown assays

For detection of βII-spectrin/αII-spectrin complexes, control and mutant GFP-βIISp proteins were coupled to GFP-bound Protein-A/G magnetic beads and incubated with lysates from HEK293 cells expressing mCherry-αIISp in IP buffer overnight at 4°C. Beads complexes were washed sequentially with IP buffer, followed by washes in TBS containing 400 mM NaCl, and TBS. Proteins were eluted in 5x PAGE loading buffer and analyzed by SDS-PAGE and western blot.

### Actin co-sedimentation assay

Interaction between purified full-length βII-spectrin proteins and actin was evaluated using the Actin Binding Protein Spin-Down Biochem Kit (#BK001, Cytoskeleton) following the manufacturer’s recommendations. In brief, full-length βII-spectrin (1 mg/ml) and α-actinin (20 mg/ml, positive control) were prepared in general actin buffer (5 mM Tris-HCl pH 8.0 and 0.2 mM CaCh) and centrifuged at 150,000 x g for 1 h at 4°C. F-actin (1 mg/ml) was prepared by incubation of purified actin in general actin buffer for 30 min on ice followed by the actin polymerization step in actin polymerization buffer (50 mM KCl, 2 mM MgCl2, 1 mM ATP) for 1 hour at 24°C. F-actin (21 μM) was incubated with either βII-spectrin (10 μM), α-actinin (2 μM), or BSA (2 μM, negative control) for 30 min at 24°C. F-actin-protein complexes were pelleted by ultracentrifugation at 150,000 x g for 1.5 h at 24°C. The presence of F-actin together with interacting proteins was assessed in the supernatant and pellet fractions by SDS-PAGE and Coomassie blue stain.

### Fluorescence image acquisition and image analysis

Confocal microscope images were taken using a Zeiss LSM780 using 405-, 488-, 561-, and 633-nm lasers. Single images and Z-stacks with optical sections of 1 μm intervals and tile scans were collected using the x10 (0.4 NA) and x40 oil (1.3 NA) objective lens. Images were processed, and measurements taken and analyzed, using Zeiss Zen, Volocity (Perkin Elmer), or NIH ImageJ software. Three-dimensional rendering of confocal Z-stacks was performed using Imaris (Bitplane).

### Time-lapse video microscopy and movie analyses

Live microscopy of neuronal cultures was carried out using a Zeiss 780 laser scanning confocal microscope (Zeiss) equipped with a GaAsP detector and a temperature- and CO2-controlled incubation chamber. Movies were taken in the mid-axon and captured at a rate of 1 frame/second for time intervals ranging from 60-300 seconds with a 40x oil objective (1.4NA) using the zoom and definite focus functions. Movies were processed and analyzed using ImageJ (http://rsb.info.nih.gov/ij). Kymographs were obtained using the KymoToolBox plugin for ImageJ (https://github.com/fabricecordelieres/IJ_KymoToolBox). In details, space (x axis in μm) and time (y axis in sec) calibrated kymographs were generated from video files. In addition, the KymoToolBox plugin was used to manually follow a subset of particles from each kymograph and report the tracked particles on the original kymograph and video files using a color code for movement directionality (red for anterograde, green for retrograde and blue for stationary particles). Quantitative analyses were performed manually by following the trajectories of individual particles to calculate dynamic parameters including, net and directional velocities and net and directional run length, as well as time of pause or movement in a direction of transport. Anterograde and retrograde motile vesicles were defined as particles showing a net displacement >3 μm in one direction. Stationary vesicles were defined as particles with a net displacement <2 μm.

### Statistical analysis

GraphPad Prism (GraphPad Software) was used for statistical analysis. Two groups of measurements were compared by unpaired Student’s t test. Multiple groups were compared by one-way ANOVA followed by a Dunnett’s multiple comparisons test.

### Molecular modeling of *SPTBN1* Variants

We used the closed conformation of utrophin CH1-CH2 closed dimer (PDB 1qag)^39^ as a template for the analogous βII-spectrin conformation to estimate its electrostatic surface profile. Molecular structures from the 6.9 Å cryo-EM structure of the CH1 actin-binding domain of Plll-spectrin bound to F-actin (PDB ID 6anu)^41^ and the structure of the CH2 domain of βII-spectrin (PDB ID 1bkr)^40^ were used for protein-protein docking predictions. The ClusPro protein-protein docking webserver^42,43^ was used to 1) dock the CH1 domain of spectrin onto F-actin, 2) dock the CH2 domain of spectrin onto F-actin, and 3) dock the CH2 domain of spectrin onto the CH1 domain of spectrin. The CH1 structure used for the dockings reported here was the model of the CH1 domain of Plll-spectrin from 6anu (chain a)^41^. This CH1 model was built based on the crystal structure of plectin (PDB ID 1mb8)^63^ by I-TASSER^64^. The CH1 domain of Plll-spectrin shares 95% sequence identity with the CH1 domain of βII-spectrin. The actin model corresponded to chains A-F of 6anu, which in turn was generated from the cryo-EM structure of actin (PDB ID 5jlh)^65^. The molecular structure of the CH2 domain of βII-spectrin from 1bkr was of a 1.1 A crystal structure^40^.

To identify the inactive closed conformation of the tandem domain (CH1-CH2) of βII-spectrin, the CH2 domain of βII-spectrin was docked onto the CH1 domain of βII-spectrin using the ClusPro webserver. The top 15 docking poses for each of the four scoring algorithms were evaluated for the placement of βII-spectrin residue L250 from the CH2 domain at the interface of the CH2/CH1 closed conformation. The top docking pose in the electrostatic scoring algorithm corresponded to a pose with a deeply buried L250 at the interface of the CH1/CH2 complex. The mutation of the equivalent residue in pill-spectrin (L253P) might disrupt the closed structure and drive the spectrin ensemble to a more open state suitable for binding to actin^40^. This same docking pose was also a top docking pose (pose 4) within the set of poses calculated by the balanced scoring algorithm. This pose was used for evaluation of the βII-spectrin mutants. It was also the same pose compared to the actinin-4 (PDB ID 6oa6) (unpublished) and utrophin (PDB ID 1qag)^39^ closed conformations.

For each of the three ClusPro protein docking analyses, the webserver provided up to 30 docking poses for each of four scoring algorithms (balanced; electrostatic-favored; hydrophobic-favored; VdW+Elec). The top 15 poses from each of the four scoring algorithms were included in the final analysis. For the dockings of the CH1 and CH2 domains from βII-spectrin onto F-actin, several of the top docking poses were to the ends of the actin segment defined as the receptor. These docking poses were immediately rejected as other actin molecules would be binding at those locations in F-actin and these sites would not be available for binding to spectrin. For CH1 docking onto F-actin, the remaining poses within the top eight docking poses predicted by the balanced and electrostatic scoring algorithms almost all corresponded to the location and orientation of CH1 molecules on actin as defined by the cryo-EM structure 6anu. For CH2 docking onto F-actin, the remaining poses within the top 8 docking poses predicted by the balanced and electrostatic scoring algorithms almost all corresponded to symmetry-related locations and poses on the F-actin. In addition, the predicted orientation of the CH2 molecules on F-actin was consistent with the known binding site of the CH1 domains, as judged by the length of the linker that would be required to join the C-terminus of the docked CH1 domain to the N-terminus of the docked CH2 domain.

βII-spectrin is a large multi-domain protein that requires a different approach for each type of domain. The SR have relatively low sequence identity to each other, and only a few have been experimentally solved, requiring independent models to be generated for each. We used RaptorX^66^ homology modeling to generate each model and assembled them into a linear conformation using Discovery Studio [Dassault Systèmes BIOVIA, Discovery Studio Modeling Environment, Release 2019, San Diego: Dassault Systèmes. 2019]. We calculated protein electrostatics using APBS^67^ and visualized structures using PyMOL [The PyMOL Molecular Graphics System, Version 2.0.7 Schrödinger, LLC.]. Individual spectrin repeats were also superimposed onto each other using a geometric algorithm^68^ as implemented in PyMOL, to investigate patterns across the fold.

### Behavioral assessment

#### Animals

Because the *Sptbn1^flox/flox^*/Nestin-Cre (βII-SpKO) mice have early mortality (typically between PND30 and PND40), testing in these mice was conducted late in the juvenile period. Subjects were 15 wildtype *(Sptbn1^flox/flox^/+*, βII-SpWT) and 5 βII-SpKO mice, taken from 5 litters. βII-SpKO mice were evaluated in two tests: open field (at PD 28-31) and acoustic startle (at PD 29-32). *Sptbn1^flox/flox^/Nestin-Cre* (PlI-SpHet), which have normal survival rates, were subjected to a more expansive battery of tests. βII-SpHet mice (n=12 per genotype, all males) underwent the following tests, with order planned so that more stressful procedures occurred closer to the end of the study.

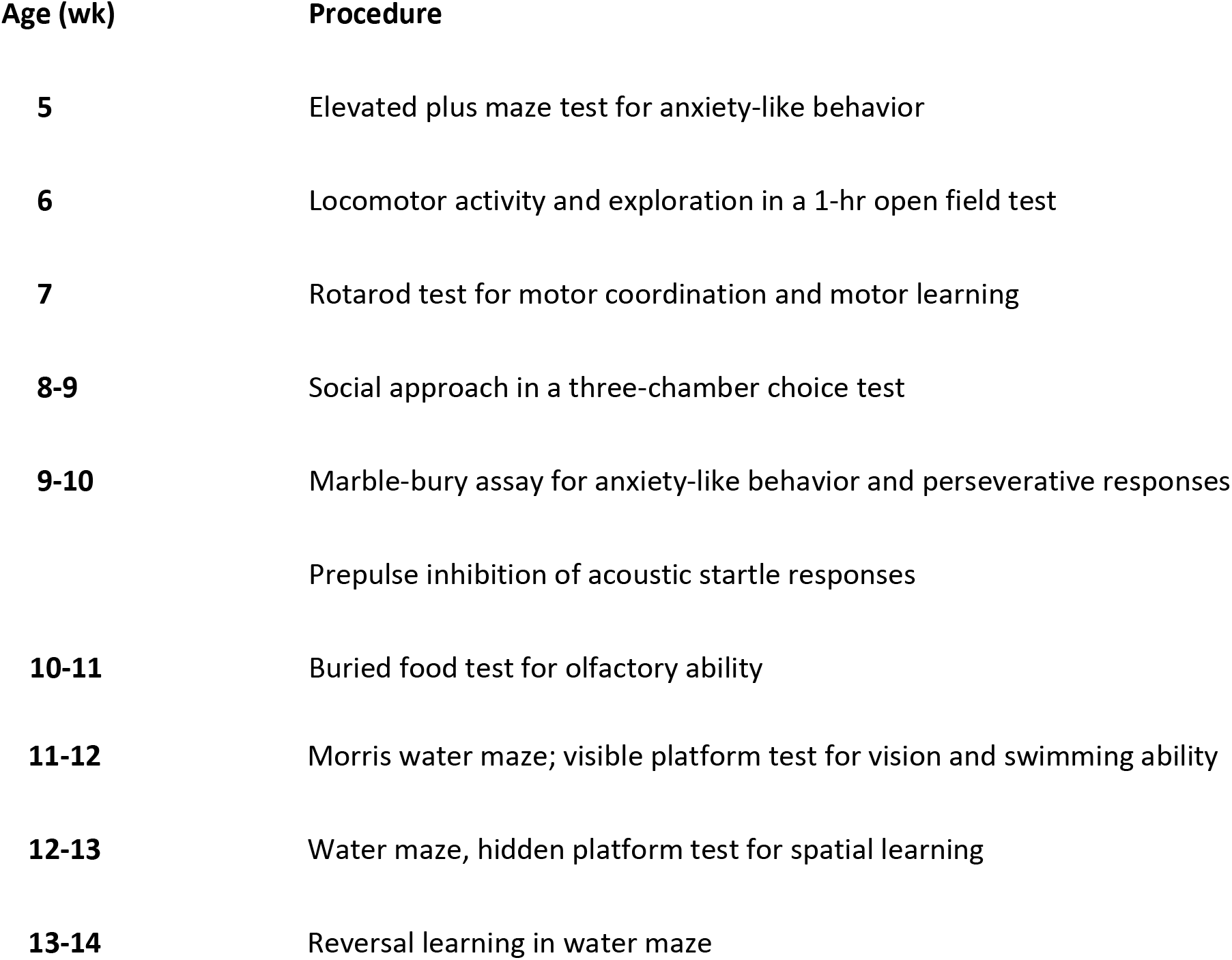

#### Elevated plus maze

A five-min test for anxiety-like behavior was carried out on the plus maze (elevation, 50 cm H; open arms, 30 cm L; closed arms, 30 cm L, walls, 20 cm H). Mice were placed in the center (8 cm × 8 cm) at the beginning of the test. Measures were taken of percent open arm time and open arm entries, and total number of arm entries.

#### Open field

Exploratory activity was evaluated by a 1-hr test (30-min for βII-SpKO mice) in a novel open field chamber (41 cm × 41 cm × 30 cm) crossed by a grid of photobeams (VersaMax system, AccuScan Instruments). Counts were taken of photobeam breaks in 5-min intervals, with separate measures for locomotor activity (total distance traveled) and vertical rearing movements. Anxiety-like behavior was assessed by measures of time spent in the center region.

#### Accelerating rotarod

Mice were first given three trials on the rotarod (Ugo Basile, Stoelting Co.), with 45 seconds between each trial. Two additional trials were conducted 48 hr later, to evaluate consolidation of motor learning. Rpm (revolutions per minute) progressively increased from 3 to a maximum of 30 rpm. across five minutes (the maximum trial length), and latency to fall from the top of the rotating barrel was recorded.

#### Social approach in a three-chamber choice test

Mice were evaluated for the effects of *Sptbn1* deficiency on social preference. The procedure had three 10-minute phases: habituation, sociability, and social novelty preference. In the sociability phase, mice were presented with a choice between proximity to an unfamiliar C57BL/6J adult male (“stranger 1”), versus an empty cage. In the social novelty phase, mice were presented with the already-investigated stranger 1 and a new unfamiliar mouse (“stranger 2”). The test was carried out in a rectangular, three-chambered Plexiglas box (60 cm L, 41. 5 cm W, 20 cm H). An automated image tracking system (Noldus Ethovision) provided measures of time in spent within 5 cm proximity to each cage and entries into each side of the social test box.

#### Marble-burying

Mice were tested for exploratory digging in a Plexiglas cage, placed inside a sound-attenuating chamber with ceiling light and fan. The cage floor had 5 cm of corncob bedding, with 20 black glass marbles (14 mm diameter) set up in a 5 × 4 grid on top of the bedding. Measures were taken of the number of marbles buried by the end of the 30-min test.

#### Buried food test

Mice were presented with an unfamiliar food (Froot Loops, Kellogg Co.) in the home cage several days before the test. All home cage food was removed 16-24 hr before the test. The assay was conducted in a tub cage (46 cm L, 23.5 cm W, 20 cm H), containing paper chip bedding (3 cm deep). One Froot Loop was buried in the cage bedding, and mice were given 15 min to locate the buried food. Latency to find the food was recorded.

#### Acoustic startle

This procedure was used to assess auditory function, reactivity to environmental stimuli, and sensorimotor gating. The test was based on the reflexive whole-body flinch, or startle response, that follows exposure to a sudden noise. Mice were evaluated for startle magnitude and prepulse inhibition, which occurs when a weak prestimulus leads to a reduced startle in response to a subsequent louder noise. Startle amplitudes were measured by force displacement of a piezoelectric transducer (SR-Lab, San Diego Instruments). The test had 42 trials (7 of each type): no-stimulus trials, trials with the acoustic startle stimulus (40 msec; 120 dB) alone, and trials in which a prepulse stimulus (20 msec; either 74, 78, 82, 86, or 90 dB) occurred 100 msec before the onset of the startle stimulus. Levels of prepulse inhibition at each prepulse sound level were calculated as 100 - [(response amplitude for prepulse stimulus and startle stimulus together/response amplitude for startle stimulus alone) x 100].

#### Morris water maze

The water maze (diameter = 122 cm) was used to assess spatial and reversal learning, swimming ability, and vision. The procedure had three phases: visible platform, acquisition in the hidden platform task, and reversal learning (with the platform moved to a new location). For each phase, mice were given 4 60-sec trials per day. Measures were taken of time to find the escape platform (diameter = 12 cm) and swimming velocity by an automated tracking system (Noldus Ethovision). Criterion for learning was an average group latency of 15 sec or less to locate the platform. At the end of the acquisition and reversal phases, mice were given a one-min probe trial in the maze without the platform. Selective quadrant search was evaluated by measuring number of crosses over the location where the platform (the target) had been placed during training, versus the corresponding areas in the other three quadrants.

### Statistical Analyses for behavioral tests

All testing was conducted by experimenters blinded to mouse genotype. Statview (SAS, Cary, NC) was used for data analyses. One-way or repeated measures analysis of variance (ANOVA) were used to determine effects of genotype. Post-hoc analyses were conducted using Fisher’s Protected Least Significant Difference (PLSD) tests only when a significant F value was found in the ANOVA. For all comparisons, significance was set at p<0.05.

## Data Availability

All original data files are available upon request

## Web Resources

Genome Aggregation Database (GnomAD), https://gnomad.broadinstitute.org/

ClinVar, https://www.ncbi.nlm.nih.gov/clinvar/

Combined Annotation Dependent Depletion (CADD), https://cadd.gs.washington.edu/

Mutation Taster, http://www.mutationtaster.org/

PolyPhen2, http://genetics.bwh.harvard.edu/pph2/

Protein Variation Effect Analyzer (PROVEAN), http://provean.jcvi.org/index.php

Sorting Intolerant from Tolerant (SIFT), https://sift.bii.a-star.edu.sg/

PredictSNP2, http://loschmidt.chemi.muni.cz/predictsnp2/

M-CAP, http://bejerano.stanford.edu/mcap/

## Acknowledgments

We thank all the families who participated in this study. We thank Drs. Matthew Rasband and Klaus-Armin Nave for the gift of the βII-spectrin conditional null and the Nex-cre mice, respectively. We thank Dr. Natallia V. Riddick for her assistance with the behavioral studies. We thank Drs. Beverly Koller and Karen Mohlke for their insightful comments on this manuscript and Dr. James Bear for helpful discussions. M.A.C, L.E.S-R., and E.W.K. were supported by the Center for Individualized Medicine at Mayo Clinic. D.N.L. was supported by the University of North Carolina at Chapel Hill (UNC-CH) School of Medicine as a Simmons Scholar, by the National Ataxia Foundation, and by the US National Institutes of Health (NIH) grant R01NS110810. Microscopy was performed at the UNC-CH Neuroscience Microscopy Core Facility, supported, in part, by funding from the NIH-NINDS Neuroscience Center Grant P30 NS045892 and the NIH-NICHD Intellectual and Developmental Disabilities Research Center Support Grant U54 HD079124, which also supported, in part, the behavioral studies. Research reported in this manuscript was supported by the NIH Common Fund, through the Office of Strategic Coordination/Office of the NIH Director under Award Number U01HG007672 (Duke University to Dr. Vandana Shashi). The content is solely the responsibility of the authors and does not necessarily represent the official views of the NIH. This research was made possible through access to the data and findings generated by the 100,000 Genomes Project. The 100,000 Genomes Project is managed by Genomics England Limited (a wholly owned company of the Department of Health and Social Care). The 100,000 Genomes Project is funded by the National Institute for Health Research and NHS England. The Wellcome Trust, Cancer Research UK and the Medical Research Council have also funded research infrastructure. The 100,000 Genomes Project uses data provided by patients and collected by the National Health Service as part of their care and support.

## Author Contributions

M.A.C. and D.N.L. conceived and planned the study with input from Q.K.T and R.C.S. M.A.C. managed the collection, analysis, and interpretation of patient clinical data with Q.K.T., R.C.S., and D.N.L. D.N.L. designed the cell biology, histology, and biochemistry studies, performed these with K. A. B., B. A. C., D. A., and S. D., and analyzed the data. S.T., M.T.Z., B.T. and D.N.L. performed the structural modeling. K.M.H. and S.M. performed the mouse behavioral studies. M.C.S. contributed reagents. M.A.C. and D.N.L. wrote the manuscript with contributions from R.C.S., S.M, M.T.Z, and B.T. E.W.K. and D.N.L. supervised the study. All other authors including Q.K.T. and R.C.S. contributed clinical data. All authors approved the final manuscript.

